# Comparison of infection control strategies to reduce COVID-19 outbreaks in homeless shelters in the United States: a simulation study

**DOI:** 10.1101/2020.09.28.20203166

**Authors:** Lloyd A.C. Chapman, Margot Kushel, Sarah N. Cox, Ashley Scarborough, Caroline Cawley, Trang Nguyen, Isabel Rodriguez-Barraquer, Bryan Greenhouse, Elizabeth Imbert, Nathan C. Lo

## Abstract

**Background:** COVID-19 outbreaks have occurred in homeless shelters across the US, highlighting an urgent need to identify the most effective infection control strategy to prevent future outbreaks.

**Methods:** We developed a microsimulation model of SARS-CoV-2 transmission in a homeless shelter and calibrated it to data from cross-sectional polymerase-chain-reaction (PCR) surveys conducted during COVID-19 outbreaks in five shelters in three US cities from March 28 to April 10, 2020. We estimated the probability of averting a COVID-19 outbreak when an exposed individual is introduced into a representative homeless shelter of 250 residents and 50 staff over 30 days under different infection control strategies, including daily symptom-based screening, twice-weekly PCR testing and universal mask wearing.

**Results:** The proportion of PCR-positive residents and staff at the shelters with observed outbreaks ranged from 2.6% to 51.6%, which translated to basic reproduction number (*R*_0_) estimates of 2.9–6.2. The probability of averting an outbreak diminished with higher transmissibility (*R*_0_) within the simulated shelter and increasing incidence in the local community. With moderate community incidence (~30 confirmed cases/1,000,000 people/day), the estimated probabilities of averting an outbreak in a low-risk (*R*_0_=1.5), moderate-risk (*R*_0_=2.9), and high-risk (*R*_0_=6.2) shelter were, respectively: 0.35, 0.13 and 0.04 for daily symptom-based screening; 0.53, 0.20, and 0.09 for twice-weekly PCR testing; 0.62, 0.27 and 0.08 for universal masking; and 0.74, 0.42 and 0.19 for these strategies combined.

**Conclusions:** In high-risk homeless shelter environments and locations with high community incidence of COVID-19, even intensive infection control strategies (incorporating daily symptom-screening, frequent PCR testing and universal mask wearing) are unlikely to prevent outbreaks, suggesting a need for non-congregate housing arrangements for people experiencing homelessness. In lower-risk environments, combined interventions should be employed to reduce outbreak risk.

## Introduction

The Coronavirus disease 2019 (COVID-19) pandemic caused by infection with severe acute respiratory syndrome coronavirus 2 (SARS-CoV-2) poses great risk to people experiencing homelessness. Across the United States (US), the estimated 568,000 people who experience homelessness nightly [1] are likely to suffer a disproportionate disease burden and need for hospitalization [2,3]. People experiencing homelessness are on average older and have a high prevalence of comorbidities that are risk factors for severe COVID-19 [2]. Multiple outbreaks in homeless shelters have occurred in several cities including San Francisco, Boston, Seattle and Atlanta with attack rates of up to 67% [4–7]. Homeless shelters have remained open in many cities despite high incidence of infection in the community, concern about the risk of further outbreaks, and uncertainty over the effectiveness of different infection control strategies. There is an immediate need to identify the best infection control strategy to reduce the risk of outbreaks and assess the safety of continuing to operate congregate shelters where transmission in the community is high.

The role of shelters and associated infection control practices in transmission of COVID-19 among people experiencing homelessness is still poorly understood. Given current understanding that SARS-CoV-2 virus is transmitted predominantly through respiratory droplets, with some airborne and fomite transmission [8], there is a need to consider policies to limit transmission within high-density congregate living environments. Different infection control strategies are currently recommended based on the level of transmission in the external community [9]. These include routine symptom screening, polymerase chain reaction (PCR) testing, universal mask wearing, and relocation of individuals at high risk of severe disease to non-congregate settings [10]. There is limited evidence on the effectiveness of strategies to reduce transmission in congregate settings, and thus further research is urgently needed to guide city-level policy across the US.

The goal of this study is to identify the most effective infection control strategy to slow the spread of COVID-19 among people experiencing homelessness who reside in shelters. We address this pressing question by estimating comparative health outcomes of key infection control strategies using a simulation model calibrated to data on homeless shelter outbreaks.

## Methods

### Microsimulation model

We developed an individual-level stochastic susceptible-exposed-infectious-recovered (SEIR) model [11] to simulate transmission of SARS-CoV-2 in a congregate shelter population (Additional file 1: Figure S1). The model defines individuals as susceptible, exposed, infectious, or immune to SARS-CoV-2 (Additional file 1: Table S1). We constructed the model to include important aspects of the natural history of COVID-19, including sub-clinical infection, pre-symptomatic transmission, and age-specific differences in risk of severe symptoms (see Additional file 1 for full details). In the model, susceptible individuals become infected with SARS-CoV-2 at a rate proportional to the prevalence of infectious individuals inside the shelter and their infectiousness (assuming homogeneous mixing), plus a static force of infection based on the background infection incidence in the community outside the shelter. Upon infection, individuals enter a latent infection stage in which they incubate the virus but are not infectious. They then progress to become infectious and contribute to ongoing transmission. An age-dependent fraction of infected individuals develop clinical symptoms with associated risk of hospitalization and death (Additional file 1: Table S2), while the remainder have sub-clinical infection. Individuals who recover from infection are assumed to remain immune.

### Data

The model was calibrated using aggregate data from PCR testing conducted during COVID-19 outbreaks in five shelters in three US cities – San Francisco (n=1), Boston (n=1) and Seattle (n=3) [4,6,7] – from March 28–April 10, 2020. We obtained de-identified individual-level data from the outbreak in the San Francisco shelter (see Additional file 1 and Additional file 1: Table S3 for details), which is fully described elsewhere [5]. As of April 10, 2020, a total of 89 individuals (84 residents, 5 staff) of 175 tested (130 residents, 45 staff) in the shelter were PCR-positive. We obtained aggregate data from the outbreaks in the Boston and Seattle shelters, where identified COVID-19 cases triggered mass testing events [4,6,7]. In the Boston shelter, 147 of 408 residents and 15 of 50 staff were PCR-positive during testing conducted April 2–3, 2020. The numbers of residents and staff tested and positive in the three Seattle shelters (shelters A, B and C) at two testing events conducted March 30–April 1 and April 7–8, 2020 are given in Additional file 1: Table S4. For the San Francisco shelter, we used daily census data to inform the shelter population size, which decayed over time, and risk stratification for disease severity by age and comorbidity status (Additional file 1: Figure S2). For the other shelters, we assumed a constant population size over time.

### Model calibration

We calibrated the model to the aggregate numbers of individuals PCR-positive out of those tested in each shelter (daily data for the San Francisco shelter, cross-sectional for the Seattle and Boston shelters) using approximate Bayesian computation techniques (see Additional file 1). We fitted the following parameters: (i) the basic reproduction number *R*_0_ (the average number of secondary infections generated by the average infectious individual in an entirely susceptible shelter population), (ii) the number of latently infected individuals who initially entered the shelter *E*_0_, and (iii) the number of days before the first case was identified that these individuals entered the shelter *D* (Table 1). The remaining parameters were sourced from literature on natural history and epidemiology of SARS-CoV-2 (Table 1 and Additional file 1: Table S5).

**Table 1.**
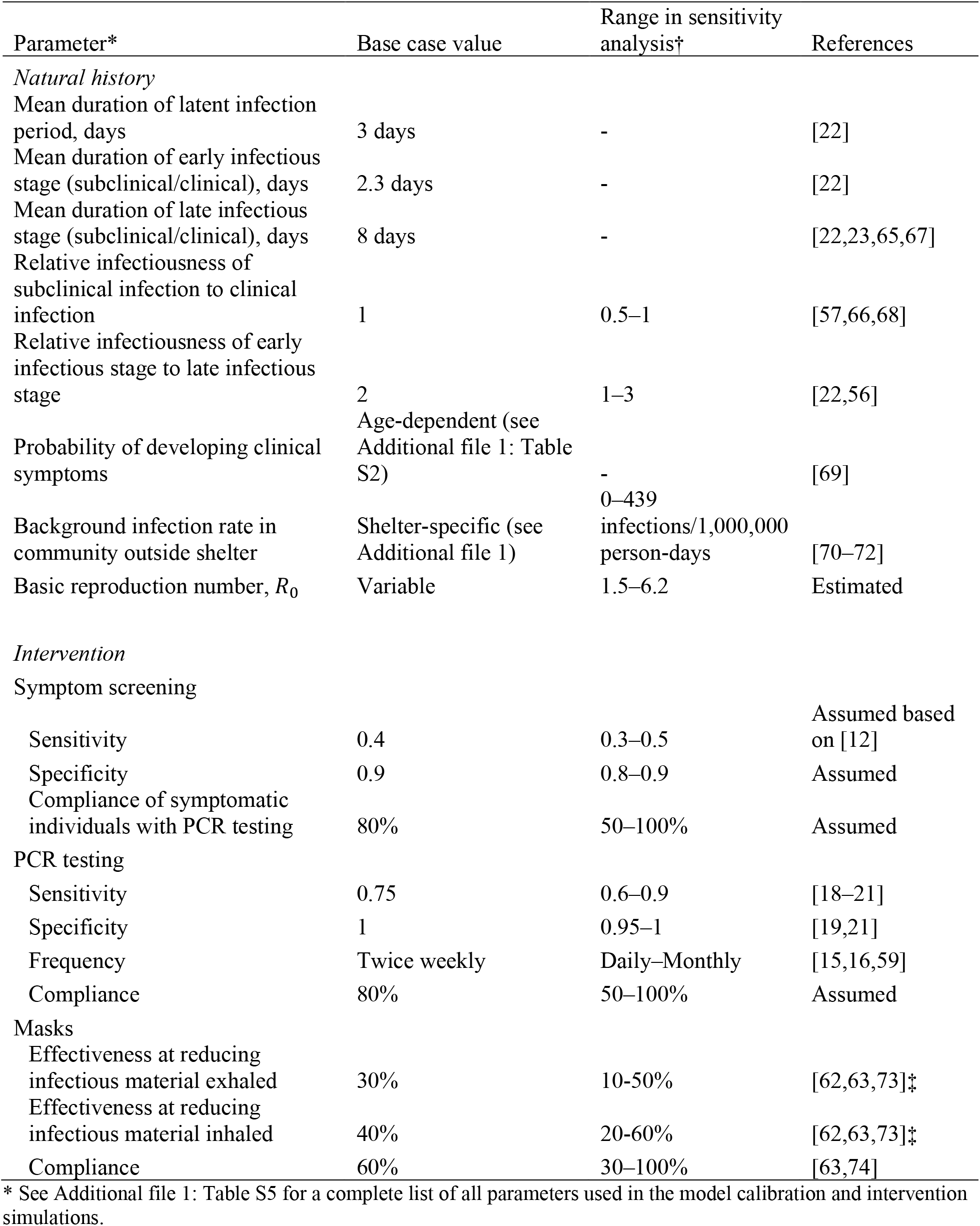
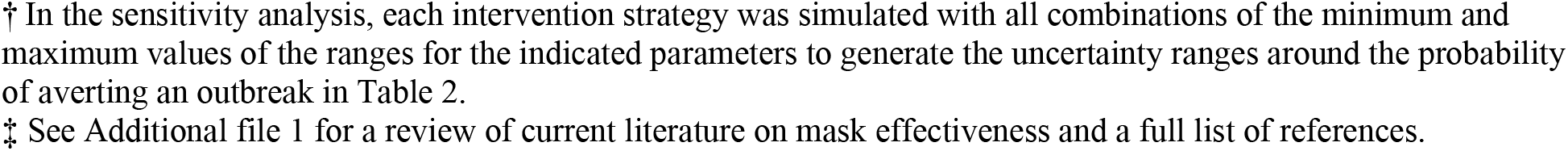
Microsimulation input parameters based on observed outbreak data from homeless shelters in Seattle, Boston and San Francisco.

| Parameter* | Base case value | Range in sensitivity analysis† | References |
| --- | --- | --- | --- |
| *Natural history* |  |  |  |
| Mean duration of latent infection period, days | 3 days | - | [22] |
| Mean duration of early infectious stage (subclinical/clinical), days | 2.3 days | - | [22] |
| Mean duration of late infectious stage (subclinical/clinical), days | 8 days | - | [22,23,65,67] |
| Relative infectiousness of subclinical infection to clinical infection | 1 | 0.5–1 | [57,66,68] |
| Relative infectiousness of early infectious stage to late infectious stage | 2 | 1–3 | [22,56] |
| Probability of developing clinical symptoms | Age-dependent (see Additional file 1: Table S2) | - | [69] |
| Background infection rate in community outside shelter | Shelter-specific (see Additional file 1) | 0–439 infections/1,000,000 person-days | [70–72] |
| Basic reproduction number, $R_0$ | Variable | 1.5–6.2 | Estimated |
| *Intervention* |  |  |  |
| Symptom screening |  |  |  |
| Sensitivity | 0.4 | 0.3–0.5 | Assumed based on [12] |
| Specificity | 0.9 | 0.8–0.9 | Assumed |
| Compliance of symptomatic individuals with PCR testing | 80% | 50–100% | Assumed |
| PCR testing |  |  |  |
| Sensitivity | 0.75 | 0.6–0.9 | [18–21] |
| Specificity | 1 | 0.95–1 | [19,21] |
| Frequency | Twice weekly | Daily–Monthly | [15,16,59] |
| Compliance | 80% | 50–100% | Assumed |
| Masks |  |  |  |
| Effectiveness at reducing infectious material exhaled | 30% | 10–50% | [62,63,73]‡ |
| Effectiveness at reducing infectious material inhaled | 40% | 20–60% | [62,63,73]‡ |
| Compliance | 60% | 30–100% | [63,74] |

\* See Additional file 1: Table S5 for a complete list of all parameters used in the model calibration and intervention simulations.
† In the sensitivity analysis, each intervention strategy was simulated with all combinations of the minimum and maximum values of the ranges for the indicated parameters to generate the uncertainty ranges around the probability of averting an outbreak in Table 2.
‡ See Additional file 1 for a review of current literature on mask effectiveness and a full list of references.

### Infection control strategies

We simulated six infection control strategies (Additional file 1: Table S6), selected via informal consultation with public health experts. 1) *Daily symptom-based screening*: daily screening of all individuals in the shelter involving a temperature and symptom survey. Individuals who screened positive were PCR tested, with 80% compliance, and isolated for 1 day pending the test result; if negative, they returned to the population. We assumed that isolated individuals were unable to transmit or become infected. We used published data on the sensitivity of symptom-based screening with time since infection [12], which suggests that close to 100% of symptomatic cases (a subset of all true cases) would eventually be detected under repeated daily screening based on the definition of being symptomatic, even with low sensitivity of symptom screening on any one occasion (here assumed to be 40% to give a 98% probability of detection after 8 days of daily symptom screening). Despite reports of low specificity of symptom screening [13,14], a high specificity of 90% was assumed to prevent unrealistic levels of PCR testing and isolation of symptom-positive individuals awaiting test results. We assumed a minimum of 3 days between repeat PCR tests for the same individual based on typical clinical practice and test turnaround times. 2) *Routine PCR testing*: twice-weekly PCR testing of residents and staff based on prior literature analyzing reduction in transmission and cost-effectiveness under different testing frequencies [15–17]. We assumed 75% sensitivity and 100% specificity of PCR testing based on published literature [18–21], a mean duration of detectable viral load (starting prior to development of symptoms) of 20 days (Additional file 1: Figure S3) [22–27], and 80% compliance with testing. We assumed test results were returned in 1 day, after which time individuals who tested PCR-positive were removed from the shelter population. 3) *Universal mask wearing*: wearing of masks by all persons within the shelter. We assumed that mask wearing reduced the amount of infectious SARS-CoV-2 material breathed into the air by infected individuals by 30% and that inhaled by susceptible individuals by 40% based on a review of the literature on filtration efficiencies of masks and the impact of mask wearing on infection risk (see Additional file 1), and that 60% of individuals adhered to mask wearing [28–31]. 4) *Relocation of “high-risk” individuals*: moving high-risk individuals (defined as those ≥60 years and/or with co-morbidities) to single hotel rooms, modelled by replacing such individuals with lower-risk individuals. 5) *Routine PCR testing of staff only*: twice-weekly testing of staff only, assuming 80% compliance. 6) *Combination strategy*: strategies 1–4 combined. Daily symptom screening (strategy 1) was included in all strategies as it is considered a minimum requirement under CDC guidelines for control practices in homeless shelters [32].

### Prediction of impact of infection control strategies

For each intervention strategy we simulated transmission within a shelter of 250 residents and 50 staff (based on an average shelter size) over 30 days starting with one latently infected individual 1000 times (to account for stochastic uncertainty). The time period was chosen to capture the trajectory of an outbreak and differential benefits of strategies. The primary outcome was the probability of averting an outbreak (defined as 3 or more infections originating within the shelter in any 14-day period [33,34]) under each strategy, with secondary outcomes of the proportional reductions in the total numbers of SARS-CoV-2 infections and clinical cases, and total numbers of hospitalizations, deaths and PCR tests used. Only individuals who tested positive were removed from the shelter population. The initial population was chosen to have the same composition in terms of proportions in different risk groups (by age and co-morbidity status) as the San Francisco shelter. We estimated the probability of averting an outbreak under each intervention strategy (compared with no interventions) for each calibrated *R*_0_ value for a range of different background infection rates estimated from incidence of confirmed cases in Seattle, Boston and San Francisco (see Additional file 1 for details). To account for potential upward bias in the estimated *R*_0_ range due to fitting to data from shelters with high attack rates, we performed the same simulations for a shelter environment with a low *R*_0_ of 1.5. The analyses were conducted in R version 4.0.0 [35] and the data and model code are available at https://github.com/LloydChapman/COVID_homeless_modelling.

### Sensitivity analysis

We conducted a multi-way sensitivity analysis to assess the impact of uncertainty in key natural history and intervention parameters – relative infectiousness of subclinical infection and the early infectious stage, sensitivities and specificities of symptom screening and PCR tests, testing and masking compliances, and mask effectiveness – on the results, by simulating each intervention strategy across all combinations of the minimum and maximum values of these parameters over their uncertainty ranges (Table 1). We explored the impact of PCR testing frequency on the probability of averting an outbreak by varying the testing frequency in strategy 2 from daily to monthly.

## Results

### Model calibration

The model reproduced the numbers of PCR-positive individuals in the cross-sectional surveys in the Seattle and Boston shelters (Additional file 1: Figure S4) and the observed numbers of PCR-positive individuals and symptomatic cases over time for the outbreak in the San Francisco shelter (Additional file 1: Figures S4–S5). The estimated *R*_0_ values ranged from 2.9 (95% CI 1.1–6.7) for Seattle shelter B to 6.2 (95% CI 4.0–7.9) for the San Francisco shelter (Additional file 1: Table S7), with corresponding estimated cumulative infection incidences at the end of the testing period of 14% (95% CI 1–41%) and 83% (95% CI 72–92%) (Additional file 1: Table S8). The median estimated number of infections initially introduced was 3 for all shelters (95% CI 1– 5), but with a relatively flat posterior distribution extending to the bounds of the uniform prior distribution, reflecting considerable uncertainty in this parameter (Additional File 1: Figure S10). The estimated date of introduction of infection ranged from 10 days (95% CI 7–14 days) before the first case was identified for Seattle shelter B to 21 days (95% CI 17–26 days) before for San Francisco.

### Impact of infection control strategies

Table 2 shows the projected impact of the six infection control strategies considered, for different transmission environments. Daily symptom screening performed poorly for all levels of transmission (probability of averting an outbreak = 0.04 for San Francisco *R*_0_= 6.2, and probability = 0.35 for *R*_0_ = 1.5). Relocating individuals at high-risk of clinical symptoms combined with symptom screening performed similarly to symptom screening alone (probability of averting an outbreak = 0.04–0.33 for *R*_0_ = 6.2–1.5). Twice-weekly PCR testing of staff provided some additional benefit over daily symptom screening at lower levels of transmission (probability of averting an outbreak = 0.04–0.41 for *R*_0_ = 6.2–1.5). Twice-weekly PCR testing of all individuals and universal masking yielded higher probabilities of averting an outbreak of 0.09–0.53 and 0.08–0.62 for *R*_0_ = 6.2–1.5. The combination strategy involving daily symptom screening, twice-weekly PCR testing of all individuals, universal masking, and removal of high-risk individuals gave the highest probability of averting an outbreak (0.19–0.74 for *R*_0_= 6.2– 1.5), but still prevented a minority of outbreaks in all but the lowest-risk setting.

**Table 2.** Probability of averting an outbreak* over a 30-day period in a generalized homeless shelter† with simulated infection control strategies.

| Infection control strategy‡ | Probability of averting an outbreak (UR)§ |  |  |  |
| --- | --- | --- | --- | --- |
| | $R_0 = 1.5$<br>(low-risk) | $R_0 = 2.9$<br>(Seattle) | $R_0 = 3.9$<br>(Boston) | $R_0 = 6.2$<br>(San Francisco) |
| 1) Symptom screening | 0.35 (0.21–0.67) | 0.13 (0.05–0.39) | 0.08 (0.02–0.28) | 0.04 (0.00–0.15) |
| 2) Routine twice-weekly PCR testing | 0.53 (0.34–0.87) | 0.20 (0.10–0.64) | 0.12 (0.05–0.50) | 0.09 (0.01–0.33) |
| 3) Universal mask wearing | 0.62 (0.26–0.99) | 0.27 (0.07–0.94) | 0.19 (0.04–0.90) | 0.08 (0.01–0.77) |
| 4) Relocation of high-risk individuals | 0.33 (0.20–0.68) | 0.13 (0.05–0.40) | 0.07 (0.02–0.29) | 0.04 (0.00–0.15) |
| 5) Routine twice-weekly PCR testing of staff only | 0.41 (0.28–0.72) | 0.15 (0.07–0.40) | 0.09 (0.03–0.33) | 0.04 (0.01–0.17) |
| 6) Combination strategy | 0.74 (0.40–1) | 0.42 (0.13–0.99) | 0.29 (0.07–0.97) | 0.19 (0.02–0.91) |

UR = uncertainty range; $R_0$ = basic reproduction number.
\* Outbreak defined as $\geq 3$ infections originating within the shelter in any 14-day period.
† Generalized homeless shelter defined as 250 residents and 50 staff with a background infection rate estimated from data for Boston (~120/1,000,000 person-days).
‡ All strategies included daily symptom screening.
§ UR generated from parameter sensitivity analysis (see Table 1 and Additional file 1).

The probability of averting an outbreak under each intervention strategy decreased with increasing transmission potential (*R*_0_) inside the shelter and with increasing infection incidence in the community outside the shelter (Figure 1). Even under the combination strategy, the probability of averting an outbreak in an average-transmission-potential shelter (*R*_0_ = 2.9) decreased from 0.77 to 0.12 as the background infection rate increased from 0 to 439 cases per 1 million person-days (the estimated background infection rate in San Francisco between June 27 and July 10, 2020).

**Figure 1.**
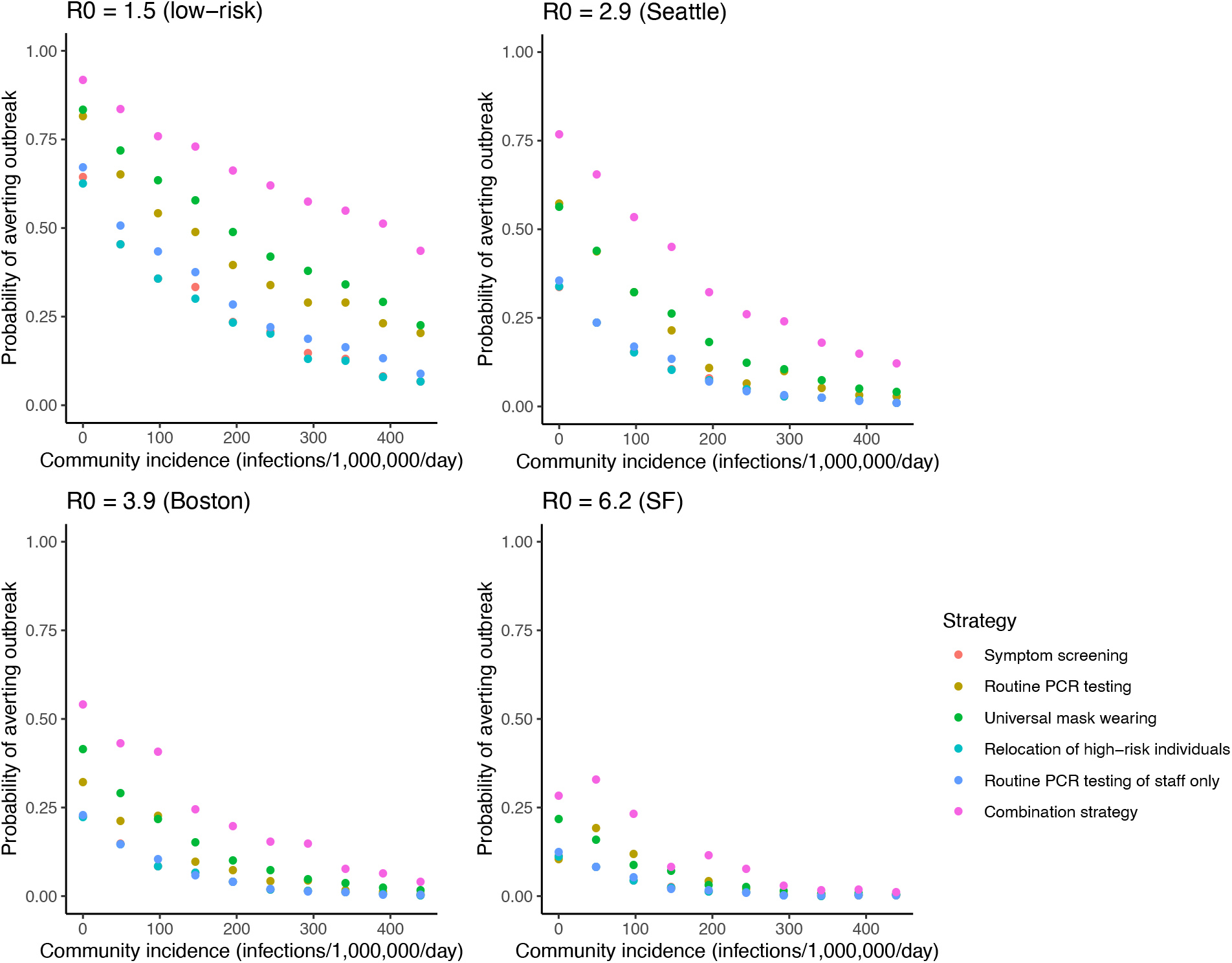
Impact of incidence of infection in the community on the probability of averting an outbreak in a generalized homeless shelter under different intervention strategies for different *R*_0_ values. The probability of averting an outbreak (≥3 infections over any 14-day period) in a generalized homeless shelter of 250 residents and 50 staff over 30 days was estimated for different infection incidences in the community using the microsimulation model described in the text. A thousand simulations of the counterfactual no-intervention scenario and each of the intervention strategies were run and the probability of averting an outbreak calculated as the proportion of simulations with an outbreak in the no-intervention scenario in which there was no outbreak in the intervention scenario. SF = San Francisco.

The relative reduction in infection incidence under the different infection control strategies followed the same pattern as the probability of averting an outbreak (Additional file 1: Table S10 and Figure 2).

**Figure 2.**
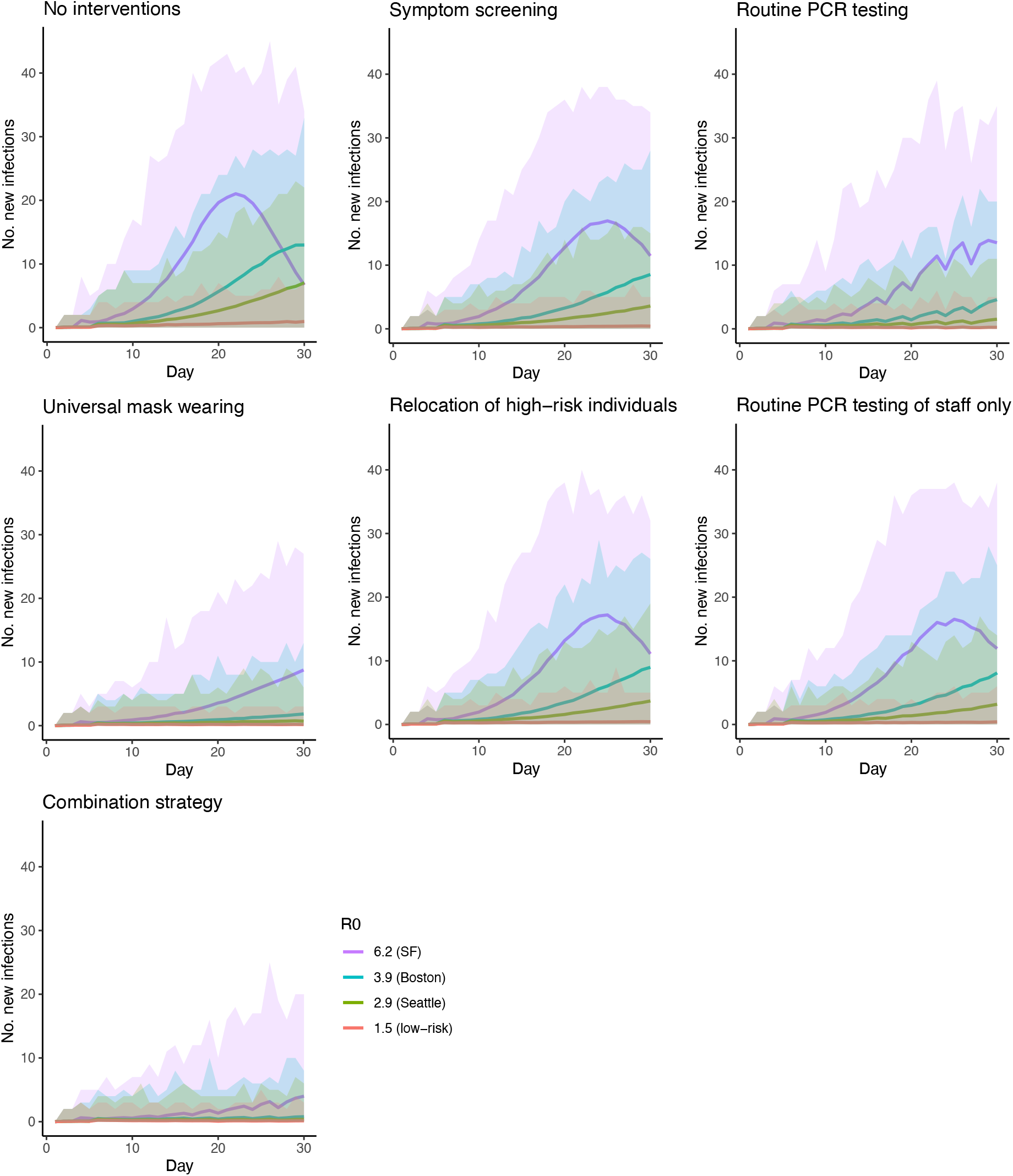
Predicted number of SARS-CoV-2 infections over a 30-day period in a generalized homeless shelter under different infection control strategies for different *R*_0_ values. Solid lines show mean daily numbers of new infections and shaded areas show minimum and maximum daily numbers over 1000 simulations. Generalized homeless shelter defined as 250 residents and 50 staff. Background infection rate in the community outside the shelter of approximately 120 infections/1,000,000 person-days. SF = San Francisco.

PCR test requirements were approximately three times higher (at an average of 6.6 tests per person per month) under twice-weekly PCR testing of all individuals than when only testing individuals identified as symptomatic in daily symptom screening (2.0 tests/person/month), and approximately two times higher than when only testing staff twice-a-week (2.8 tests/person/month) (Additional file 1: Table S11).

### Sensitivity analysis

The probability of averting an outbreak was most sensitive to uncertainty in masking compliance and effectiveness and relative infectiousness of the early infectious stage, with the mean probability of averting an outbreak under combined interventions across all combinations of the minimum and maximum values of the other parameters varying from 0.40–0.71 for 30–100% masking compliance, 0.49–0.62 and 0.48–0.63 for 10–50% and 20–60% mask exhalation and inhalation effectiveness, and 0.63–0.48 for early-stage relative infectiousness of 1–3 for *R*_0_= 2.9 (Additional file 1: Figure S9). After this, the probability of averting an outbreak was most sensitive to PCR sensitivity and testing compliance, with the mean probability of averting an outbreak under combined interventions varying from 0.50–0.61 and 0.51–0.60 over the uncertainty ranges of these parameters. Decreasing the frequency of PCR testing from daily to monthly decreased the probability of averting an outbreak for *R*_0_= 1.5, 2.9 and 3.9 from 0.71 to 0.33, 0.28 to 0.12, and 0.21 to 0.08 respectively, but had little impact on the already low probability of averting an outbreak for *R*_0_ = 6.2 (Figure 3).

**Figure 3.**
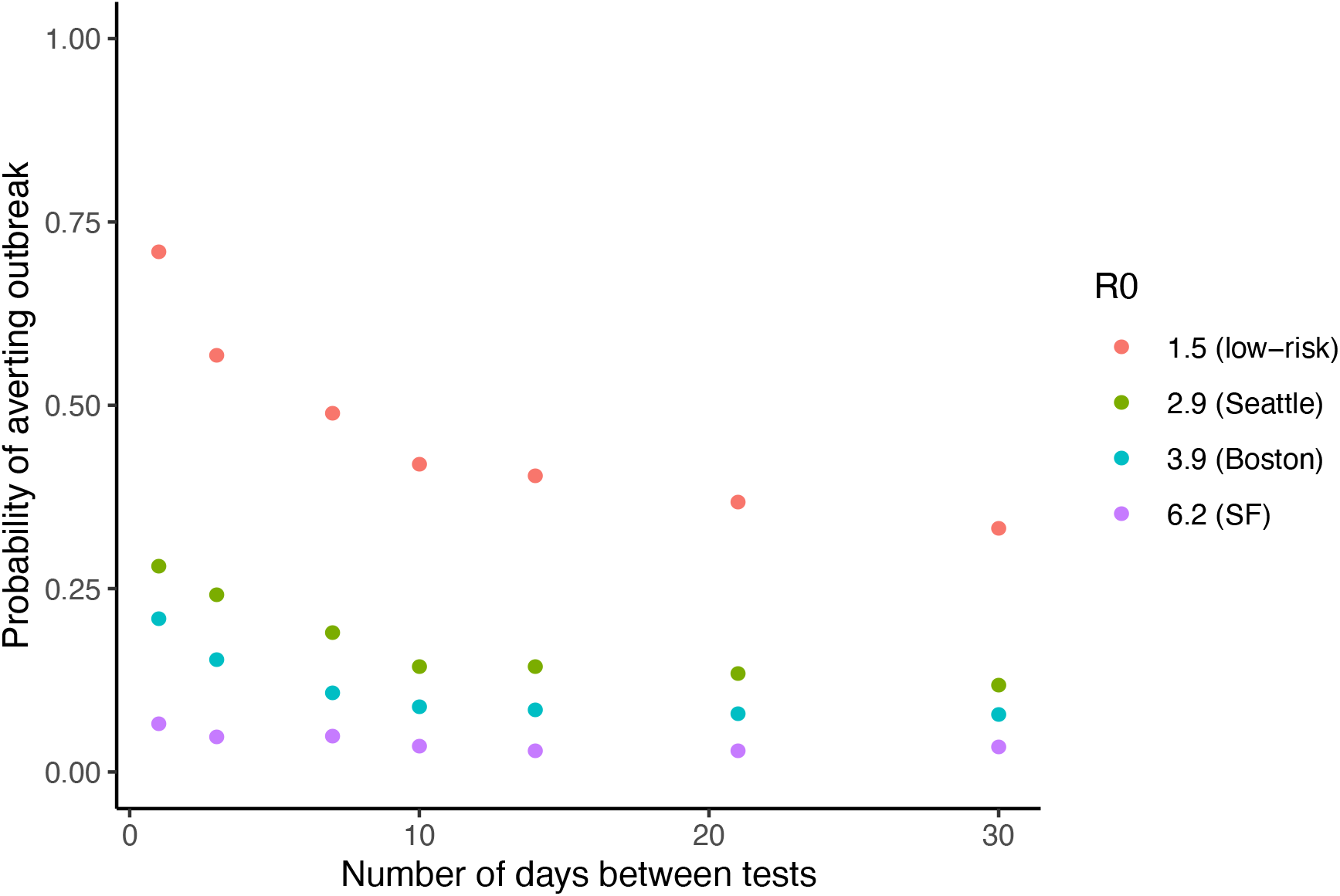
Impact of varying the frequency of routine PCR testing of residents and staff on the probability of averting an outbreak in a generalized homeless shelter for different *R*_0_ values. The probability of averting an outbreak (≥3 infections over any 14-day period) over 30 days was estimated for different frequencies of routine PCR testing from daily (1 day between tests) to monthly (30 days between tests). Generalized homeless shelter defined as 250 residents and 50 staff. Background infection rate in the local community of approximately 120 infections/1,000,000 person-days. SF = San Francisco.

## Discussion

Several outbreaks of COVID-19 with high attack rates have occurred in homeless shelters across the US, and there remains uncertainty over the best infection control strategies to reduce outbreak risk in shelters. In this study, we applied a simulation analysis to identify infection control strategies to prevent future outbreaks. We found that in high-risk shelters that are unable to maximize basic infection control practices that sufficiently reduce the transmissibility of SARS-CoV-2 (e.g. social distancing, reduced living density), no additional infection control strategy is likely to prevent outbreaks. Similarly, in cities with high community incidence, no infection control practices are likely to prevent an outbreak. In contrast, in lower-risk shelters with low background community incidence, the implementation of strategies such as symptom screening, routine PCR testing, and masking would help reduce outbreak risk.

We found a wide range of transmissibility of SARS-CoV-2 based on observed outbreaks in homeless shelters, which greatly affects intervention impact. We estimated basic reproduction numbers (*R*_0_) of 2.9–6.2 from aggregate PCR test data from outbreaks in five shelters in Seattle, Boston and San Francisco between March and April 2020. This range of *R*_0_ values is at the high end of estimates reported in the literature [36–39], and likely reflects a high degree of heterogeneity in infectiousness between individuals [38,40–43] and a highly conducive environment for transmission within these shelters due to lack of existing infection control practices and high living density at the time of the outbreaks. For these *R*_0_ values and representative background infection rates, we found that the infection control strategies considered are unlikely to prevent outbreaks (probability < 50%), even when combined. Nevertheless, they do reduce incidence of infection and clinical disease and slow the growth of the outbreak (Figure 2). Our *R*_0_ estimates are likely not entirely representative of general transmission potential in shelters now given that the outbreaks occurred early during the pandemic when control measures were limited, and that non-outbreaks and smaller outbreaks may go undetected or unreported. Control measures such as rehousing of individuals to single hotel rooms appear to have been successful and incidence has in general been lower in the homeless population than anticipated [44]. However, there have been subsequent large outbreaks in homeless shelters despite reduced shelter density and stringent control efforts [45–47]. This supports our finding that outbreaks in congregate shelters remain likely even with fairly intensive infection control practices.

In lower transmissibility settings, e.g. with *R*_0_ = 1.5, which may be more representative of typical shelters now due to improved social distancing and basic infection control practices, the intervention strategies we have considered are more likely to prevent outbreaks (probability up to nearly 75% under combined interventions, for a moderate background infection rate of approximately 120/1,000,000/day).

A key remaining issue given limited availability of alternative housing for people experiencing homelessness is identifying the characteristics that distinguish low-risk shelters (those similar to the *R*_0_ = 1.5 scenario considered here) that can be operated with low outbreak risk with implementation of infection control strategies. Data are limited, but available evidence suggests that social distancing and reductions in super-spreading are likely to be key factors [38,40,48– 50]. Strategies that may achieve these goals include reducing living density, spacing bedding, reducing communal activities, and adopting staffing models that limit social contacts.

The fact that intervention impact and the probability of averting an outbreak decrease significantly with increasing background infection rate in the community (Figure 1) suggests a need for alternative housing arrangements for people experiencing homelessness in locations in which community incidence is moderate to high – 100–500 infections/1,000,000/day, equivalent to 25–125 confirmed cases/1,000,000/day assuming four-fold underreporting (see Additional file 1). In lower background incidence settings, combined daily symptom-based screening, twice-weekly PCR testing, universal masking and relocation of high-risk individuals to non-congregate settings would reduce outbreak risk, and limit incidence of infection and severe disease if outbreaks do occur.

Our findings broadly agree with those of two other modelling studies of interventions against COVID-19 in homeless shelters: one in the US [51], the other in England [52]. The former found that a combination of daily symptom screening with PCR testing of symptom-positive individuals, universal PCR testing every 2 weeks, and alternative care sites for those with mild/moderate COVID-19 would significantly reduce infections, while remaining cost effective, but unlike our analysis did not consider variation in the effectiveness of interventions with community incidence. The latter study supports our results on the high risk of outbreaks in congregate homeless shelters, as it found that outbreaks in homeless shelters were likely even when incidence in the general population is low, and estimated that closure of congregate shelters during the first pandemic wave in England averted over 90% of infections.

Each infection control strategy is limited in some aspect [22,53–57]. Symptom-based screening has very low sensitivity to detect infections early in the clinical course (when people are most infectious), and poor specificity [12–14,58]. The impact of routine PCR testing is limited by imperfect PCR sensitivity (~75%), especially early in the infection course [19], as well as need for frequent testing and missing onset of infectiousness between testing periods. Other analyses support our finding that testing less than once or twice weekly leaves a high risk of outbreaks (e.g. testing once every two weeks gives a 30% lower probability of averting an outbreak than twice-weekly testing, Figure 3) [15,16,59]. However, once- or twice-weekly testing may be financially and logistically infeasible. Similarly, relocation of high-risk persons to independent housing is resource intensive. Frequent testing and universal masking also suffer issues with adherence, and may not be possible for all individuals at all times in homeless shelters.

This study has a number of limitations. Due to limited data availability, we only calibrated the model to a small number of shelter outbreaks, the *R*_0_ estimates for which are likely to be higher than for the average shelter since they occurred early in the pandemic and larger outbreaks are more likely to be reported. The cross-sectional aggregate nature of the majority of the data also led to wide uncertainty intervals around the fitted parameters, without independent identifiability between them (Additional file 1: Figure S10). Our results suggest that universal masking would significantly reduce the risk of outbreaks in homeless shelters, even with 60% compliance. However, the impact of masking is highly sensitive to the assumed masking effectiveness and compliance, estimates for which still vary considerably despite accumulating evidence that masks reduce infection risk [60–64]. Many uncertainties in the biology of SARS-CoV-2 transmission remain, particularly regarding differential infectiousness over time and by severity of illness, and the relationship of PCR positivity and infectiousness [22,65,66]. Our assumption of equal infectiousness for different individuals means that our model is unlikely to fully reproduce super-spreading events [38,40]. We made several simplifying assumptions in modelling transmission within the shelter and from the surrounding community, namely: homogenous mixing within the shelter population, no entry of new people, a stable background infection rate over time and full immunity upon recovery from infection given the short duration of the simulation. Our assumption that individuals who are isolated within homeless shelters while awaiting test results are unable to transmit or become infected may have led to slight overestimation of the impact of testing, since in reality isolation is not perfect. We assumed homogeneous mixing due to a lack of contact data for the shelter outbreaks, which meant that we were not able to consider cohorting and contact tracing as interventions.

This study defines conditions for operating homeless shelters with lower risk of COVID-19 outbreaks and estimates the impact of various interventions on outbreak risk. Our findings demonstrate the need for combined interventions (symptom-based screening, PCR testing, and masking) and regular testing to protect persons experiencing homelessness from COVID-19, while highlighting the limitations of these interventions in preventing outbreaks.

## Data Availability

All data and code used for the analysis are available at: https://github.com/LloydChapman/COVID_homeless_modelling

## Abbreviations

CI: credible interval
COVID-19: Coronavirus disease 2019
PCR: polymerase chain reaction
SARS-CoV-2: Severe acute respiratory syndrome coronavirus 2
UR: uncertainty range
US: United States

## Ethics approval and consent to participate

This study was considered exempt non-human subject research based on use of de-identified secondary data by the University of California, San Francisco Institutional Review Board.

## Consent for publication

Not applicable.

## Availability of data and materials

The datasets generated and/or analyzed during the current study are available online at https://github.com/LloydChapman/COVID_homeless_modelling.

## Competing interests

The authors declare that they have no competing interests. NCL has received funding from the World Health Organization for unrelated work.

## Funding

This work was supported by the University of California, San Francisco (UCSF) and the UCSF Benioff Homelessness and Housing Initiative. The funders had no role in the study design, data collection and analysis, preparation of the manuscript, or the decision to submit the manuscript for publication.

## Authors’ contributions

NCL and MK conceived the study. LACC and NCL designed and conducted the analysis and drafted the manuscript. SNC, AS, CC, TN, EI acquired and curated the data from the San Francisco shelter outbreak. NCL and MK acquired funding. All authors contributed to, read, and approved the final manuscript. LACC and NCL had full access to all of the data in the study and take responsibility for the integrity of the data and the accuracy of the data analysis.

## Acknowledgements

We sincerely appreciate the hard work and public health efforts of all those involved in the collection of the data used in this article. LACC would like to thank Renata Retkute, Amanda Minter, Simon Spencer and TJ McKinley for helpful discussions regarding Approximate Bayesian Computation.

## Supporting information

**Additional file 1 (pdf): Supplementary methods and results:** Details of the model and model calibration. **Table S1:** Definition of states in the transmission model. **Table S2:** Risk of clinical symptoms and hospitalization by age group and co-morbidity status. **Table S3:** Numbers of PCR-positive individuals by day of test result and daily new symptom onsets in San Francisco shelter March 28–April 10, 2020. **Table S4:** Numbers of residents and staff PCR tested and PCR positive at three shelters in Seattle during two testing events March 30–April 1, 2020 and April 7–8, 2020. **Table S5:** Input parameters for microsimulation of COVID-19 transmission in homeless shelters. **Table S6:** Different intervention strategies tested. **Table S7:** Estimated epidemiologic parameters based on observed outbreak data from homeless shelters in Seattle, Boston and San Francisco. **Table S8:** Estimated cumulative infection incidence at the end of the PCR testing period in homeless shelters in Seattle, Boston and San Francisco. **Table S9:** Probability of averting an outbreak over a 30-day period in a generalized homeless shelter with simulated infection control strategies for different background infection rates in the community outside the shelter. **Table S10:** Reductions in the total number of infections and symptomatic cases over a 30-day period in a generalized homeless shelter with simulated infection control strategies for different background infection rates in the community outside the shelter. **Table S11:** Numbers of PCR tests used under each infection control strategy. **Figure S1:** Structure of stochastic individual-level susceptible-exposed-infectious-recovered (𝒮-ℰ-ℐ-ℛ) model of COVID-19 transmission in homeless shelter. **Figure S2:** Daily numbers of residents by risk group present in the San Francisco shelter March 29–April 10, 2020. **Figure S3:** Distribution of duration of detectable viral load from start of late infectious stage. **Figure S4:** Calibration of microsimulation to observed PCR testing data from outbreaks in homeless shelters in Seattle, Boston and San Francisco. **Figure S5:** Calibration of microsimulation to additional data from San Francisco shelter outbreak. **Figure S6:** Outbreak size distributions 30 days after introduction of infection in a generalized homeless shelter under different infection control strategies for *R*_0_= 1.5 (low-risk setting). **Figure S7:** Outbreak size distributions 30 days after introduction of infection in a generalized homeless shelter under different infection control strategies for *R*_0_ = 2.9 (Seattle). **Figure S8:** Outbreak size distributions 30 days after introduction of infection in a generalized homeless shelter under different infection control strategies for *R*_0_ = 6.2 (San Francisco). **Figure S9:** Spider diagrams showing the sensitivity of the estimated probability of averting an outbreak to variation in key natural history and intervention parameters for different *R*_0_ values. **Figure S10:** Posterior distributions and pairwise correlation plots for calibrated model parameters – *R*_0_, *E*_0_ and *T* – for (A)-(C) Seattle shelters A–C, (D) Boston shelter and (E) San Francisco shelter

## Notes

**Conflicts of Interest:** The authors declare no conflicts of interest.

### Competing Interest Statement

The authors have declared no competing interest.

