## Additional file 1 for "Comparison of infection control strategies to reduce COVID-19 outbreaks in homeless shelters in the United States: a simulation study"

### Supplementary methods

#### Model description

Figure S1 shows the structure of the discrete-time stochastic individual-level susceptible-exposed-infectious-recovered (SEIR) model used to simulate transmission in the shelter. On any given day $t$ each individual is in one of the seven states shown in the flow diagram and defined in Table S1.

The probabilities that individual $i$ is infected on day $t$ or avoids infection on day $t$ given that they are susceptible to infection are:

$$p_i(t) \ = 1 - e^{-\lambda_i(t)}, \quad i \in \mathcal{S}(t)$$

$$1 - p_i(t) = e^{-\lambda_i(t)}, \quad i \in \mathcal{S}(t)$$

where $\lambda_i(t)$ is the force of infection on each individual $i$ on day $t$. The force of infection on each individual on each day is equal to the force of infection they are exposed to when inside the shelter, which is proportional to the prevalence of infectious individuals inside the shelter on that day and the infectiousness of these individuals, plus a background force of infection they are exposed to when outside the shelter in the community:

$$\lambda_i(t) = \frac{\beta \mathbb{I}_i(t)\left(h\left(\alpha I_{s,1}(t) + I_{s,2}(t)\right) + \alpha I_{c,1}(t) + I_{c,2}(t)\right)}{\sum_j \mathbb{I}_j(t)} + \epsilon. \qquad (1)$$

Here $\beta$ is the transmission rate coefficient within the shelter (assumed constant); $\mathbb{I}_i(t)$ is an indicator function for whether the individual is present in the shelter on day $t$; the $I(t)$'s represent the number of infectious individuals inside the shelter on day $t$ in different states of infection – subclinical and clinical, denoted by subscripts $s$ and $c$ respectively, and early and late stage, denoted by subscripts $1$ and $2$ respectively; $h$ is the infectiousness of subclinically infected individuals relative to those with clinical symptoms; $\alpha$ is the infectiousness of the early infectious stage relative to the late stage; and $\epsilon$ is the transmission rate outside of the shelter. Mixing of infectious and susceptible individuals in the shelter is assumed to be homogeneous due to a lack of contact data from the shelter outbreaks with which to parameterize inhomogeneous mixing. Transmission from the external community is modeled assuming homogeneous mixing of the shelter residents and staff with the community outside of the shelter during each day and negligible impact of infected individuals entering the community from the shelter on the overall background transmission rate. The background transmission rate is treated as constant given the relatively short durations of the outbreaks, and is estimated from the incidence of confirmed COVID-19 cases for the city of each shelter with adjustments for reporting delay, infection-to-onset time and relative risk of infection for homeless individuals as described below (see *Estimation of background infection rate* below).

We note that the formulation of the force of infection in equation (1) corresponds to frequency-dependent transmission, i.e. assumes that the number of "contacts" per infectious individual in the shelter per day is approximately constant regardless of the number of individuals present in the shelter. We make this assumption in common with other authors [1] because it is believed that the main mode of SARS-CoV-2 transmission is from person to person via respiratory droplets containing virus particles [2], i.e. occurs over short distances predominantly among close contacts of infectious individuals.

The duration $D_E$ of the latent infection stage $\mathcal{E}$, is assumed to be negative-binomial, with mean $\mu_E = 3$ days and shape parameter $r_E = 4$, i.e. probability mass function:

$$\mathbb{P}(D_E = d) = \frac{\Gamma(d + r_E)}{\Gamma(r_E)d!}\left(\frac{r_E}{r_E + \mu_E - 1}\right)^{r_E}\left(\frac{\mu_E - 1}{r_E + \mu_E - 1}\right)^{d-1}, \qquad d = 1,2,\ldots,$$

where $\Gamma(\mathrm{z}) = \int_0^\infty x^{z-1}e^{-x}dx$ is the gamma function. After passing throught the latent stage, individuals enter an early (presymptomatic) infectious stage ($\mathcal{I}_{c,1}$) leading to clinical symptoms with age-dependent probability $\rho(a_i)$, where $a_i$ is the age group (<60 years/≥60 years) of individual $i$, or an early infectious stage ($\mathcal{I}_{s,1}$) leading to subclinical infection (no symptoms/very mild symptoms) with probability $1 - \rho(a_i)$. After a negative-binomial number of days ($NB(r_1 = 4, \mu_1 = 2.3)$) of early-stage infectiousness individuals progress to late-stage subclinical ($\mathcal{I}_{s,2}$) or clinical ($\mathcal{I}_{c,2}$) infectiousness.

Following a further negative-binomially-distributed duration ($NB(r_2 = 4, \mu_2 = 8)$ with mean 8 days) subclinical cases recover and are no longer infectious and clinical cases either recover or are hospitalized, and therefore no longer contribute to transmission in the shelter ($\mathcal{R}$). See Table S1 for the key attributes of each infection state and Table S5 for a full list of model parameters and their values. The probability of hospitalization for clinical cases is age- and co-morbidity dependent (Table S2), and hospitalized cases are assumed to have a 26.1% risk of requiring intensive care based on data from Wuhan, China [3]. Cases admitted to the intensive care unit (ICU) have an age- and co-morbidity-dependent risk of death estimated from ICU data from Wuhan, China [4,5].

*Basic reproduction number, $R_0$*

The basic reproduction number for the model, defined as the average number of secondary infections caused by the average infectious individual in an entirely susceptible shelter population (in the absence of interventions) can be calculated from first principles as:

$$\begin{aligned} R_0 &= \text{probability of infection given "contact"} \times \text{"contact" rate} \times \text{duration of infectiousness} \\ &= \frac{\beta \sum_i \mathbb{I}_i(0)\left(\left(1 - \rho(a_i)\right)h + \rho(a_i)\right)(\alpha\mu_1 + \mu_2)}{\sum_i \mathbb{I}_i(0)} \end{aligned}$$

where "contact" is defined as susceptible individuals coming into contact with infectious material from infected individuals, and the sums are over all individuals in the shelter (residents and staff). Variation in the number of secondary infections per infectious individual is modeled through the variation in the durations of the early and late infectious stages described above.

*Relative infectiousness of early infectious stage and subclinical infection*
Evidence suggests that infectiousness of symptomatic COVID-19 cases is not constant over time, but peaks at or shortly before symptom onset, such that pre-symptomatic individuals are more infectious than symptomatic individuals [6,7]. We approximated this variation in infectiousness over time by treating individuals' infectiousness as constant during each of the early and late infectious stages, but higher during the early infectious stage. The relative infectiousness of the early infectious stage to the late infectious stage, $\alpha = 2$, was chosen to approximately match the estimates of the proportion of pre-symptomatic transmission of He et al [6] and the lower range of those of Casey et al [7] of 44% and 34% respectively, assuming a mean duration of the early infectious stage of $\mu_1 = 2.3$ days [6]:

$$\text{proportion of transmission from early infectious stage} = \frac{\alpha\mu_1}{\alpha\mu_1 + \mu_2} = \frac{2 \times 2.3}{2 \times 2.3 + 8} = 37\%,$$

We used the lower range of the estimates of Casey et al for the base case analysis, as we believe that estimates of pre-symptomatic infectiousness are likely to be biased upward due to behavior changes (e.g. self-isolation) upon symptom onset that reduce transmission from symptomatic individuals in general settings but are less likely to apply in congregate shelter settings. However, we also considered higher relative infectiousness (up to 3) in the sensitivity analysis. We assumed the same relative infectiousness of early-stage infection for clinical infection and subclinical infection.

Subclinically infected individuals were assumed to be as infectious as clinical cases in the base case analysis, due to limited data on the relative infectiousness of subclinical infection and the detection of similar viral load in asymptomatic individuals as symptomatic individuals in several studies [6,8]. Lower relative infectiousness of subclinical infection (down to 50%) was considered in the sensitivity analysis.

*Duration of detectable viral load*
Studies that have measured the viral load of individuals infected with SARS-CoV-2 over time since symptom onset suggest that the virus remains detectable from throat and nasal swabs and sputum and stool samples for longer (~20 days after symptom onset) than individuals remain infectious (~7 or 8 days after symptom onset) [6,9]. We therefore modelled the duration of detectable viral load for each infected individual by assigning a random draw from a truncated discretized normal distribution that characterizes the variation in this duration (Figure S3) to each individual when they enter the late infectious stages $\mathcal{I}_{s,2}$ and $\mathcal{I}_{c,2}$. We chose the parameters of the distribution based on data on the variation in times after symptom onset at which individuals' viral

loads reach the PCR detection limit from several studies [6,9–13], with minimum and maximum durations of 5 days and 37 days.

We assumed that individuals in the early infectious stages $\mathcal{I}_{s,1}$ and $\mathcal{I}_{c,1}$ always have detectable viral loads and that in the latent infection stage the viral load is undetectable. We treated the duration of detectable viral load as being the same for subclinical and clinical infection and independent of the individual's duration of infectiousness. This means that individuals can still have a detectable viral load when they are no longer infectious and are in the recovered compartment ($\mathcal{R}$). It is likely that there is some correlation between viral load and symptom severity and infectious duration (and other factors such as age), but as of yet there is insufficient data with which to parameterize these relationships and studies measuring viral loads over time by age and severity have shown mixed results [6,14–16].

*Sensitivity and specificity of PCR tests*

Although there is some evidence to suggest that the sensitivity of PCR tests varies with time since infection (i.e. that it is lower during early and late infection) [17], the data currently available to accurately characterize this variation is very limited. We therefore made the simplifying assumption that the sensitivity of PCR tests is constant with time since the start of infectiousness (time of entering the early infectious stages $\mathcal{I}_{s,1}$ and $\mathcal{I}_{c,1}$) and use a fixed sensitivity of 75% based on available data [17–20] for the base case analysis. We assumed perfect specificity of PCR tests for the base case analysis, i.e. no false positive test results, as current evidence suggests that they have high specificity (~99%) [17,20], but also considered lower specificity in the sensitivity analysis.

*Effectiveness and impact of masking*

The impact of mask wearing on transmission depends on the effectiveness with which masks reduce infectious material exhaled by infectious individuals, $\mathrm{e}_{ex}$, and that with which they reduce infectious material inhaled by susceptible individuals, $\mathrm{e}_{in}$, and compliance with mask wearing, $c$. We modeled the impact of masking via a reduction in the transmission rate coefficient:

$$\beta_m = \beta(1 - c\mathrm{e}_{in})(1 - c\mathrm{e}_{ex})$$

where $\beta_m$ is the transmission rate coefficient under "universal" masking and we have assumed that compliance with masking is the same among infectious individuals as among susceptible individuals.

We reviewed several different sources of evidence on the effectiveness of masking for reducing transmission of SARS-CoV-2. These included five systematic reviews and meta-analyses of impact on infection rates [21–25], a meta-analysis of impact of non-medical masking on infection rates in a community setting [26], a living rapid review [27], a narrative review [28], and a number of primary analyses on impact of mask wearing on infection among contacts of confirmed COVID-19 cases [29,30] and on the filtration efficiency of different types of masks [31–35]. Overall, these studies suggest that mask wearing reduces transmission of SARS-CoV-2. However, there is considerable variation in the estimated impact on infection rates and very few studies have been performed specifically for SARS-CoV-2 [21,23]. Those that have have tended to be in healthcare settings, where the estimated impact of mask wearing has generally been found to be higher than in non-healthcare settings [21–23]. Estimates of overall odds ratios (ORs) for risk of infection with respiratory viruses (SARS, MERS, influenza, H1N1, SARS-CoV-2) for mask wearers compared to non-mask wearers from meta-analyses range from 0.15 to 0.94, and 0.2-0.3 in healthcare settings and 0.53-0.56 in non-healthcare settings [21–25]. A retrospective cohort study of transmission of SARS-CoV-2 among household contacts of confirmed cases in China estimated an adjusted OR for risk of infection of 0.21 (95% CI 0.06-0.79) in households in which masks were used by at least 1 household member before the index case developed symptoms, but found no association between mask use after illness onset and infection risk [29]. Importantly, evidence from randomized control trials (RCTs) of mask effectiveness for preventing SARS-CoV-2 transmission is lacking. The only published RCT for SARS-CoV-2 to date did not find a statistically significant effect on infection risk [36], and RCTs for other respiratory viruses have either found only a weak effect of mask wearing on infection risk or non-statistically-significant associations [24,25,28].

Compliance with masking has been shown to be a crucial factor for effectiveness. A retrospective case-control study among contacts of COVID-19 patients in Thailand found that wearing a mask all the time decreased the risk of infection compared to not wearing a mask (adjusted OR 0.23, 95% CI 0.09-0.60), but that occasional mask use was not associated with a statistically significant decrease in infection risk (adjusted OR 0.87, 95% CI 0.41-1.84) [30]. A meta-analysis of 40 studies of effectiveness of masks for preventing transmission of respiratory viruses that controlled for virus type, setting, mask type, and comparison group, among other variables, found that non-medical masking reduced risk of infection in the general population by 40% (95% credible interval 20-54%) [26]. Mask material has also been shown to be an important factor for mask effectiveness, with N95 respirators being more effective at filtering droplets of the size that carry virus particles than surgical masks, and surgical masks being more effective than cloth masks [24]. Estimates of filtration efficiency for different mask types vary considerably, however, from 3 to 90% for cloth masks, to 53-96% for surgical masks, to 90-95% for N95 respirators [24,31,34,35].

To ensure that the overall reduction in transmission from masking is consistent with reductions estimated from observational studies [26], we conservatively assume in the base case analysis that masks worn in homeless shelters are 30% effective at filtering infectious particles exhaled by infected individuals and 40% effective at blocking infectious particles from being inhaled by susceptible individuals. However, we also consider a wide range of different efficacies in sensitivity analysis (10-50% and 20-60% respectively).

Data on mask use among people experiencing homelessness is lacking. Data from large surveys of US adults shows that mask use has increased over time, but that the percentages of individuals who report always wearing a mask in public and when visiting family and friends have only reached approximately 60% and 40% respectively

[26,37]. People experiencing homelessness are more likely to have issues with access to masks. It also may not be possible for them to wear a mask at all times when inside a shelter, e.g. when sleeping at night or if they have medical conditions that prevent them from wearing a mask. We therefore assume that average masking compliance inside the shelter is 60% in the base case analysis (corresponding to a 38% overall reduction in transmission with the above base case mask exhalation and inhalation efficacies), and consider compliances of 30-100% in sensitivity analysis (corresponding to a 10-80% overall reduction in transmission with the above uncertainty ranges for the efficacies).

*Estimation of background infection rate during shelter outbreaks*
We used publicly available data on daily numbers of new confirmed COVID-19 cases in the city of each shelter from county public health departments to estimate the background infection rate [38–40]. We assumed a fixed delay from infection to reporting of 7 days, corresponding to 2 days of pre-symptomatic infection and 5 days of symptoms before reporting [41,42], and so used case counts 7 days ahead as an estimate of the number of new reported infections on each day. We estimated the infection incidence in the community outside the shelter during the period of each shelter outbreak, $i_a$, from the delay-adjusted case counts for the city of the shelter over the 3 weeks prior to the end date of data collection, $T_{end}$, as:

$$i_a = \frac{\gamma \sum_{t=T_{end}-14}^{T_{end}+7} n_t}{P}, \qquad (2)$$

where $n_t$ is the number of new confirmed cases reported on day $t$, $P$ is the city population [43], and $\gamma$ is an under-reporting factor to account for only a proportion of infections being reported. For the period of the shelter outbreaks we used $\gamma = 10$, based on estimated infection-to-reported-case ratios from seroprevalence data for the period of late March–early May from 10 different localities across the US [44]. For the intervention simulations, we used $\gamma = 4$, based on lower estimates from seroprevalence data for late May-June from a continuation of the same study [45]. We then estimated the background infection rate in homeless individuals outside of the shelter as:

$$\epsilon = r i_a,$$

where $r$ is a relative risk of infection for homeless individuals. Based on data from Seattle & King County, WA, where 1.5% of the population is homeless (11199/753675) [43,46] and homeless individuals account for 2.5% (445/18130) of confirmed COVID-19 cases [38,47], we use an approximate relative risk for homeless individuals of $r = 2$.

**Model calibration**
We calibrated the model by fitting to data on numbers of PCR-positive and negative individuals in testing conducted in outbreaks in 5 homeless shelters in 3 different cities: San Francisco (n=1), Boston (n=1) and Seattle (n=3). Prevalence of PCR positivity among residents and staff during mass testing events in these shelters was markedly different, ranging from 2.6%–51.6%. This is likely partly due to testing being conducted

at different times after the outbreaks started, but likely also reflects differences in transmissibility due to other factors, such as variation in infectiousness between individuals, shelter living density, bed spacing, air ventilation quality, and differences in shared washing and eating facilities. We therefore fitted the model separately to the data from each of these outbreaks to estimate the basic reproduction number, $R_0$, for each setting. Given uncertainty in when the first infected individuals entered each shelter and how many of them there were, due to asymptomatic infection and incomplete detection of early cases, we also estimated the time since introduction of infection into the shelter at the end of the data collection period, $T$, and the initial number of latently infected individuals who entered the shelter, $E_0$. We assumed that the individual(s) who initially introduced infection into the shelter were all latently infected when they entered, since a large range of scenarios in which they were in a later infection stage or mixture of infection stages are covered by the flexibility in the introduction time and the initial number infected. We also assumed that following the initial introduction into the shelter any further introductions were solely as a result of residents and staff being infected when mixing with the community outside the shelter and then returning to the shelter.

Since more detailed individual-level data on PCR test results and symptom onset times for clinical cases was available for the outbreak in the San Francisco shelter, we also used the numbers of early symptomatic cases who tested PCR-positive and daily numbers of new symptom onsets when calibrating the model to this outbreak (see Figure S4 and Figure S5A). As exact dates of testing during the cross-sectional surveys in the Seattle and Boston shelters were not available, we assumed that all testing occurred on the last day of each survey. For all the outbreaks, we assumed that PCR tests took a day to be processed, such that results were returned and positive individuals removed the day following testing (except on April 10th at the San Francisco shelter, when negative individuals were removed instead).

Aggregate data (by age and co-morbidity risk group) on movement of individuals in and out of the shelter over time was only available for the San Francisco shelter, so we ensured that the population of this shelter matched that in the shelter registry (see *Demographics and movement of individuals in and out of the shelter* below) and assumed that the populations of the other shelters remained approximately constant over the period of data collection. Residents' age categories in the Seattle and Boston shelters were set according to data on the age distributions from [48] and [49]. For all shelters, staff were assumed to be all <60-years-old and at low risk of hospitalization and death.

*Approximate Bayesian Computation algorithm*

We fitted the model to the data using an approximate Bayesian computation sequential Monte Carlo (ABC-SMC) algorithm [50–52] to estimate $R_0$, $E_0$, and $T$. Since $E_0$ and $T$ are discrete parameters, we adapted the model selection algorithm of Toni et al [51,52] by replacing the model index as the discrete parameter with the discrete parameter pair $m = (E_0, T)$ (i.e. the model indices by the different possible combinations of $E_0$ and $T$). The algorithm starts by sampling pairs $m^* = (E_0^*, T^*)$ from the prior distribution $\pi(m)$, and corresponding $R_0$ values, $R_0^{**}$, from the prior distribution $\pi(R_0)$; simulating outbreaks with these parameter values (particles); and accepting those for which the simulated number of PCR-positives $D^*$ falls within a certain pre-specified tolerance $\epsilon_1$ of the observed number of PCR-positives $D$ according to a distance measure $d(\cdot)$, i.e. $d(D,D^*) \leq \epsilon_1$. A sequence of distributions (generations) is then constructed by repeating this process with a set of decreasing tolerances $\boldsymbol{\epsilon} = \{\epsilon_g\}_{g=1,2,\ldots}$, proposing $R_0$ values for each value of $m$ in each generation by perturbing the particles ($R_0$ values) specific to $m$ from the previous generation using a perturbation kernel $K(R_0|R_0^*)$. In this way, the particles in successive generations converge towards the joint posterior distribution of the parameters given the data. Pseudocode for the algorithm is as follows:

1. Set the number of generations $G$ and number of particles $N$.
2. Set the tolerance schedule $\epsilon_1 > \epsilon_2 > \cdots > \epsilon_G$. Set the generation index $g = 1$.
3. Set the particle index $i$ to 1.
4. Sample $m^*$ from the prior distribution $\pi(m)$. If $g = 1$, sample $R_0^{**}$ from the prior distribution $\pi(R_0)$. If $g > 1$, sample $R_0^*$ from the previous generation $\{R_0(m^*)_{g-1}\}$ with weights $w(m^*)_{g-1}$, and perturb the particle $R_0^*$ to obtain $R_0^{**} \sim K(R_0|R_0^*)$.
5. If $\pi(R_0^{**}) = 0$, return to step 4.
6. Run a simulation of the outbreak and PCR testing for the sampled values $(R_0^{**}, m^*)$ to generate a candidate dataset $D^{**}$.
7. If $\widehat{\mathbb{P}}(D|D^*) = \mathbb{I}\big(d(D,D^*) < \epsilon_g\big) = 0$, return to step 4.
8. Set $m_g^{(i)} = m^*$, and add $R_0^{**}$ to the population of particles $\{R_0(m^*)_g\}$ and calculate its weight as:
$$w_g^{(i)} = \begin{cases} 1, & \text{if } g = 1 \\ \dfrac{\pi(R_0^{**})}{\sum_{i;m_{g-1}=m^*} \dfrac{w_{g-1}^{(i)} K(R_0^{**}|R_{0,g-1}^{(i)})}{\mathbb{P}(m_{g-1} = m^*)}}, & \text{if } g > 1 \end{cases}$$
9. If $i < N$, set $i = i + 1$ and go to step 4.
10. Normalize the weights $w_g$ such that $\sum_{i=1}^{N} w_g^{(i)} = 1$.
11. Calculate the marginal probabilities for the combinations $m = (E_0, T)$, by summing the weights for each combination:
$$\mathbb{P}\big(m_g = m\big) = \sum_{i;m_g^{(i)}=m} w_g^{(i)}\Big(R_{0,g}^{(i)}, m_g^{(i)}\Big).$$
12. If $g < G$, set $g = g + 1$ and go to step 3.

We used $G = 10$ generations, with $N = 1000$ particles in each generation, and a normal perturbation kernel for $R_0$, $K(R_0|R_0^*) \sim N(R_0^*, \sigma^2)$, with standard deviation $\sigma = 1$. We used broad uniform prior distributions due to a lack of information to support more informative prior distributions: $R_0 \sim U(1,8)$, $m = (E_0, T) \sim U(1,5) \times U(14,30)$ (for all shelters except Seattle shelter B, for which $m \sim U(1,10) \times U(13,20)$), where the prior for $m$ is discrete with integer support. The wide bounds for the prior for $R_0$ were chosen based on the large range of basic reproduction numbers reported in the literature[41] and demonstrated potential of COVID-19 for superspreading events [53–57]. The 14-day lower bound of the prior for $T$ was chosen based on the symptom onsets of the first cases identified in each of the shelters being at least 9 days before the end of data collection and the mean incubation period being approximately 5 days, such that the first cases were unlikely to have been infected later than 14 days before the end of data collection. Thirty days was taken as the upper bound for $T$ based on it giving the earliest plausible time for introduction of infection into the shelters without earlier occurrence and detection of symptomatic cases. Lower bounds were used for the prior for $T$ for Seattle shelter B to reflect the fact that it did not report any symptomatic cases before the first mass testing event on March 30–April 1 and had a very low prevalence of infection at that survey.

We used the sum of squared differences between the numbers of PCR-positives on the testing days, $\boldsymbol{T}_T$, in the simulations, $D^*$, and those in the observed data, $D$, for the distance metric $d(D, D^*)$:

$$d(D, D^*) = \sqrt{\sum_{t \in \boldsymbol{T}_T} (D_t - D_t^*)^2}$$

For the tolerance schedule $\boldsymbol{\epsilon}^T = (\epsilon_1, \dots, \epsilon_G)$ we used regular steps decreasing from a discrepancy of twice the daily number of PCR-positives, $\epsilon_1 = \sqrt{\sum_t (2D_t)^2}$, to half the width of the exact binomial confidence interval on the number of PCR-positives on each day. For the San Francisco shelter, we used additional distance metrics, $d^S(D^S, D^{S*})$ and $d^C(D^C, D^{C*})$, and tolerance schedules, $\boldsymbol{\epsilon}^S$ and $\boldsymbol{\epsilon}^C$, for the differences in the simulated and observed numbers of PCR-positives among early symptomatic cases tested, $D^{S*}$ and $D^S$, on days $\boldsymbol{T}_S$ (3/30/20–4/7/20) and the differences in the simulated and observed daily numbers of symptom onsets, $D^{C*}$ and $D^C$:

$$d^S(D^S, D^{S*}) = \sqrt{\sum_{t \in \boldsymbol{T}_S} (D_t^S - D_t^{S*})^2}$$

$$d^C(D^C, D^{C*}) = \sqrt{\sum_{t=1}^{T} (D_t^C - D_t^{C*})^2}$$

Since the symptom onset data is less reliable than the PCR test data and potentially incomplete, we used less strict tolerances for $\boldsymbol{\epsilon}^S$ and $\boldsymbol{\epsilon}^C$, decreasing from discrepancies of twice the daily observed number of PCR-positives among early symptomatic cases and twice the daily number of new symptom onsets to 2/3 and 1.3 times the observed daily numbers respectively:

$$\boldsymbol{\epsilon}^S = (11,10,9,9,8,7,6,5,5,4)$$

$$\epsilon^C = (49,47,45,43,41,39,37,36,34,32).$$

Proposed values of $(R_0, E_0, T)$ were accepted at each generation $g$ only if all tolerances were satisfied, i.e. only if $d(D, D^*) \leq \epsilon_g$ and $d^S(D^S, D^{S*}) \leq \epsilon_g^S$ and $d^C(D^C, D^{C*}) \leq \epsilon_g^C$.

We assessed the performance of the algorithm by calculating the effective sample size (ESS) of the final generation of particles, $ESS = 1/\sum_{i=1}^{N}(w_G^{(i)})^2$, and the acceptance rate of proposed particles in each generation.

**Details of San Francisco shelter outbreak**
Full details of the outbreak in the San Franciso shelter are provided elsewhere [58]. Briefly, the data consisted of individual-level information on age, PCR test date and result, and partial data on symptom status, co-morbidity, and health outcome. The first two clinical cases identified in the shelter were confirmed on April 5, 2020 from PCR tests on April 4 and 5, 2020. The first case had symptom onset on March 31, the second on April 2. However, several individuals who later tested PCR-positive reported that they had symptom onset around these dates (Table S3 and Figure S5A). After the first cases were identified on April 4, contact tracing, symptom screening and PCR testing of symptomatic individuals was performed up to April 7. On April 8 and 9 mass testing of residents and staff was performed. As of April 10, 2020, 89 individuals out of 175 tested in the shelter were PCR-positive, of whom 65 were pre-symptomatic/symptomatic (2 unknown status), 4 required hospitalization (2 unknown outcome), and 1 died. Table S3 shows the numbers of positive test results returned from the different testing that was conducted by day of testing and number of new symptom onsets each day.

*Demographics and movement of individuals in and out of the shelter*
Even before the first cases in the shelter in San Francisco were identified, the shelter was not running at capacity (340 residents) and efforts were made to move high-risk individuals (those aged 60 and over or with co-morbidities) out of the shelter. Following identification of the first cases on April 5, progressively more and more individuals were removed from the shelter into isolation and quarantine at various sites; first those identified as close contacts and bedmates of the first cases and those suspected of being infected, then later on April 10 PCR-negative individuals, particularly high-risk individuals. Efforts were made to cohort the remaining residents into those who were PCR-positive and those with unknown COVID-status, but this was hampered by individuals returning to the shelter on the evening of April 10. The shelter was disbanded and all residents and staff were moved to isolation and quarantine sites on April 11.

According to the shelter register, there were a total of 255 residents who were present at some point from March 29 to April 10, 2020. Figure S2 shows the breakdown of the number of residents in each of the different risk groups (no co-morbidities and under-60, co-morbidities and under-60, no co-morbidities and 60 or over, co-morbidities and 60 or over) present on each day from March 29 to April 10. We initialized the resident population of the shelter in the simulations such that the numbers in the different risk groups matched those present on March 29, and assumed the numbers in the different groups remained constant prior to March 29. We assumed that the proportions of the remaining individuals not present on March 29 in the different risk groups were the same as among those present. The movement of residents in and out of the shelter each day was simulated by randomly drawing individuals from each risk group to remove/add such that the number present in each risk group matched that in the register, accounting for the removal of symptomatic PCR-positive individuals from the testing from April 4–8 and their risk group.

A total of 64 staff, covering general running of the shelter, support services, maintenance, laundry and food services over 3 shifts per day with approximately 20-25 staff on each shift, were present at some point from March 29 to April 10. Due to a lack of detailed information on staff demographics and movement we assumed that all staff had low risk of clinical symptoms, hospitalization and death (were all under-60 without co-morbidities), and were present in the shelter each day for approximately the same amount of time as the average resident, such that they had the same risk of infection as residents.

**Estimation of impact of different interventions**

We estimated the impact of six different intervention strategies, listed in Table S6 with their component interventions, on the probability of averting an outbreak and the total numbers of infections, clinical cases, hospitalizations and deaths over 30 days in a shelter of 250 residents and 50 staff into which one latently infected individual is introduced by comparing output of simulations in which there were interventions with counterfactual simulations without any interventions. An outbreak was defined as ≥3 cases that originated within the shelter within any 14-day period, which we determined by probabilistically assigning an infection source (background transmission vs infectious individuals within the shelter) to each infected individual upon infection in the simulations and tracking the number of infections whose source was within the shelter over time. We ran 1000 simulations for each intervention strategy and the counterfactual scenario, and calculated the probability of averting an outbreak from pairs of counterfactual and intervention simulations as the proportion of simulation pairs with an outbreak in the no-intervention scenario in which there was no outbreak in the intervention scenario:

$$\mathbb{P}(\text{outbreak averted}) = \mathbb{P}(\text{no outbreak with intervention}|\text{outbreak without intervention})$$
$$= \frac{\#(\text{pairs with no outbreak in intervention simulation \& an outbreak in counterfactual simulation})}{\#(\text{counterfactual simulations with an outbreak})}$$

Reductions in cumulative incidence of infections and clinical cases under each intervention strategy were calculated as the median percentage reduction in total number of infections/clinical cases between the counterfactual simulation and the intervention simulation across all simulation pairs, where the percentage reduction was treated as 0 if there were no cases in the counterfactual simulation.

**Scenario and sensitivity analyses**

To assess the effect of the transmission potential within the shelter ($R_0$) and the background infection rate ($i_a$) on intervention impact, we predicted the impact of the different intervention strategies for the different $R_0$ estimates from the calibration ($R_0 = 2.9$ (Seattle A), $3.9$ (Boston), 6.2 (San Francisco)) and $R_0 = 1.5$ (representing a lower-risk setting) for different background infection rates estimated from incidence of confirmed cases in Seattle, Boston and San Francisco. The background infection rates were estimated as in Equation (2) but with the limits in the sum replaced by July 4 and July 17, 2020 (to represent reported incidence for June 27–July 10, 2020, with a 7-day infection-to-reporting delay) and an infection-to-reported-case ratio, $\gamma$, of 4 [45]. This gave background infection rates ranging from 122/1,000,000/day for Boston to 439/1,000,000/day for San Francisco. We used the limits of this range and the mean across the three cities, along with a zero background infection rate, for the scenario analyses. The results are provided in Table 2 in the main text and Tables S9 and S10. We also assessed the variation in the probability of averting an outbreak under each intervention strategy for a larger number of background infection rates over the same range (Figure 1 in the main text).

We assessed the sensitivity of the intervention impact estimates to uncertainty in key natural history and intervention parameters (relative infectiousness of subclinical infection and the early infectious stage, sensitivities and specificities of symptom screening and PCR tests, testing and masking compliances, and mask effectiveness) by simulating each intervention strategy with all combinations of minimum and maximum values of these parameters over their uncertainty ranges (Table S5) for the base case background infection rate of 122/1,000,000/day. We then calculated the minimum and maximum values of the probability of averting an outbreak over all parameter combinations to generate uncertainty ranges around the base case estimates of the probability of averting an outbreak (Table 2 in the main text). The sensitivity of the probability of averting an outbreak under the combination strategy to variation in the different parameter values is shown in Figure S9 and discussed in the main text.

### Supplementary results

#### Model calibration

Figure S10 shows the posterior distributions and pairwise correlation plots for the calibrated parameters $R_0$, $E_0$ and $T$ for each of the shelters. The considerable uncertainty in the parameter estimates due to the predominantly cross-sectional aggregate nature of the data is reflected in the broad posterior distributions, covering most of the range of the prior distributions for the parameters for all the shelters except the San Francisco shelter, and the strong correlation between $R_0$ and $T$ for the Boston and San Francisco shelters.

The effective sample sizes of the output for the different shelters ranged between 640 for Seattle shelter C and 951 for the San Francisco shelter, indicating that a sufficient number of particles was used to estimate the posterior distributions. The acceptance rates varied across shelters and decreased over successive generations, remaining above 50% for all of the Seattle shelters but decreasing to 4% for the San Francisco shelter, but overall suggest that the algorithm sampled efficiently from the posterior distributions.

#### Impact of infection control strategies

The relative impact of the different infection control strategies on reducing cumulative infection incidence followed the same pattern as the probability of averting an outbreak (cf. Table S10 with Table 2 in the main text and Table S9). However, the percentage reduction in cumulative incidence varied non-linearly with $R_0$ due to the bimodal nature of the outbreak size distribution (Figures S6–S8), such that the highest percentage reductions were achieved for the $R_0 = 2.9$ scenario. Daily symptom screening alone, and daily symptom screening with relocation of high-risk individuals led to reductions in cumulative incidence of 7% and 6% for $R_0 = 6.2$ to 45% and 44% for $R_0 = 2.9$ (for a background infection rate of 122/1,000,000/day). Twice-weekly PCR testing of staff provided modest additional benefit, increasing the percentage reduction to 8-50%. Reductions under twice-weekly PCR testing of all residents and staff and universal masking were much greater (31–75% and 67–83%), though the impact of PCR testing attenuated more than that of masking with increasing $R_0$. The highest percentage reductions of 72–92% were achieved under the combination strategy, with the biggest gain from combining interventions occurring in the highest transmissibility setting ($R_0 = 6.2$).

The pattern of impact of the intervention strategies in terms of reduction in total numbers of clinical cases was the same (Table S10), except for relocation of high-risk individuals, which led to mostly greater percentage reductions in clinical cases than symptom screening and routine PCR testing of staff. Total numbers of hospitalizations and deaths over 30 days were small with or without interventions (medians ≤ 4) and therefore not considered relevant at the scale of a single shelter.

**Table S1. Definition of states in the transmission model**

| State | Symbol | Infectious | Symptomatic | Detectable viral load | Immune |
|---|---|---|---|---|---|
| Susceptible | $\mathcal{S}(t)$ | ✗ | ✗ | ✗ | ✗ |
| Exposed to infection | $\mathcal{E}(t)$ | ✗ | ✗ | ✗ | ✗ |
| Early subclinical infection | $\mathcal{I}_{s,1}(t)$ | ✓ | ✗ | ✓ | ✗ |
| Late subclinical infection | $\mathcal{I}_{s,2}(t)$ | ✓ | ✗/✓ (no/mild symptoms) | ✓ | ✗ |
| Early clinical infection | $\mathcal{I}_{c,1}(t)$ | ✓ | ✗ | ✓ | ✗ |
| Late clinical infection | $\mathcal{I}_{c,2}(t)$ | ✓ | ✓ | ✓ | ✗ |
| Recovered | $\mathcal{R}(t)$ | ✗ | ✗ | ✓/✗ | ✓ |

**Table S2. Risk of clinical symptoms and hospitalization by age group and co-morbidity status**

| Risk group | Probability of developing clinical symptoms, $\rho(a_i)$ | Probability of hospitalization for clinical cases | Probability of death for hospitalized cases admitted to ICU |
|---|---|---|---|
| Low risk: age <60 yrs + no co-morbidities | 0.473 | 0.040 | 0.22 |
| Moderate risk: age <60 yrs + co-morbidities | 0.473 | 0.085 | 0.33 |
| High risk: age ≥60 yrs + no co-morbidities | 0.747 | 0.289 | 0.52 |
| Very high risk: age ≥60 yrs + co-morbidities | 0.747 | 0.618 | 0.77 |

**Table S3. Numbers of PCR-positive individuals by day of test result and daily new symptom onsets in San Francisco shelter March 28–April 10, 2020**

| Date | Number tested in random testing | Number PCR-positive in random testing | Number of early symptomatic cases tested | Number of early symptomatic cases PCR-positive | Number of new symptom onsets |
|---|---|---|---|---|---|
| Mar 28 | 1 | 0 | 0 | - | 2 |
| Mar 29 | 1 | 0 | 0 | - | 0 |
| Mar 30 | 2 | 0 | 1 | 0 | 3 |
| Mar 31 | 0 | - | 0 | - | 2 |
| Apr 1 | 0 | - | 0 | - | 3 |
| Apr 2 | 0 | - | 0 | - | 1 |
| Apr 3 | 0 | - | 0 | - | 1 |
| Apr 4 | 1 | 0 | 1 | 1 | 2 |
| Apr 5 | 0 | - | 1 | 1 | 3 |
| Apr 6 | 0 | - | 3 | 2 | 7 |
| Apr 7 | 1 | 0 | 5 | 5 | 3 |
| Apr 8 | 89 | 35 | - | - | 4 |
| Apr 9 | 64 | 44 | - | - | 16 |
| Apr 10 | 5 | 1 | - | - | 15 |

**Table S4. Numbers of residents and staff PCR tested and PCR positive at three shelters in Seattle during two testing events March 30–April 1, 2020 and April 7–8, 2020**

| Shelter | Testing event 1 (Mar 30–Apr 1, 2020) | | Testing event 2 (Apr 7–8, 2020) | |
|---|---|---|---|---|
| | No. tested | No. (%) positive | No. tested | No. (%) positive |
| Seattle A* | | | | |
| Residents | 43 | 7 (16.3) | - | - |
| Staff | 15 | 4 (26.7) | - | - |
| Seattle B | | | | |
| Residents | 74 | 2 (2.7) | 52 | 4 (7.7) |
| Staff | 2 | 0 (0) | 8 | 1 (12.5) |
| Seattle C | | | | |
| Residents | 37 | 6 (16.2) | 44 | 10 (22.7) |
| Staff | 10 | 0 (0) | 7 | 1 (14.3) |

* Shelter A closed April 5, 2020, so data from testing event 2 was not used.

**Table S5. Input parameters for microsimulation of COVID-19 transmission in homeless shelters**

| Parameter | Symbol | Base case value | Range in sensitivity analysis | References |
|---|---|---|---|---|
| *Demography* | | | | |
| Number of residents | | | | |
| Seattle A | | 43 | - | [48] |
| Seattle B | | 109 | - | [48] |
| Seattle C | | 93 | - | [48] |
| Boston | | 408 | - | [59] |
| San Francisco | | Time-varying (see Figure S2) | - | |
| Number of staff | | | | |
| Seattle A | | 15 | - | [48] |
| Seattle B | | 8 | - | [48] |
| Seattle C | | 10 | - | [48] |
| Boston | | 50 | - | [59] |
| San Francisco | | 64 | - | |
| Age group of individual $i$ (<60 years, ≥60 years) | $a_i$ | Shelter-specific (see text) | | [48,49] |
| *Natural history* | | | | |
| Mean duration of latent infection period | $\mu_E$ | 3 days | - | [6] |
| Shape parameter of negative-binomially-distributed latent infection period | $r_E$ | 4 | | [1] |
| Mean duration of early infectious stage (subclinical/clinical) | $\mu_1$ | 2.3 days | - | [6] |
| Shape parameter of negative-binomially-distributed early infectious stage (subclinical/clinical) | $r_1$ | 4 | | [1] |
| Mean duration of late infectious stage (subclinical/clinical) | $\mu_2$ | 8 days | - | [6,9,60,61] |
| Shape parameter of negative-binomially-distributed late infectious stage (subclinical/clinical) | $r_2$ | 4 | | [1] |
| Relative infectiousness of subclinical infection to clinical infection | $h$ | 1 | 0.5–1 | [8,62,63] |
| Relative infectiousness of early infectious stage to late infectious stage | $\alpha$ | 2 | 1–3 | [6,7] |
| Probability of developing clinical symptoms | $\rho(a_i)$ | Age-dependent (see Table S2) | - | [1] |

| | | | | |
|---|---|---|---|---|
| Mean duration of detectable viral load from start of late infectious stage | | 20 days | - | [6,9–13] |
| Minimum duration of detectable viral load from start of late infectious stage | | 5 days | - | [6,9–13] |
| Maximum duration of detectable viral load from start of late infectious stage | | 37 days | - | [11,13] |
| Mean time from symptom onset to hospitalization | | 8 days | | Assumed same as duration of late infectious stage [41,64] |
| Probability of hospitalization for clinical cases | | Age- and co-morbidity dependent (see Table S2) | - | [4] |
| Probability of ICU admission among hospitalized cases | | 0.261 | - | [3] |
| Probability of death for hospitalized cases admitted to ICU | | Age- and co-morbidity dependent (see Table S2) | - | [4] |
| Infection-to-reported-case ratio | $\gamma$ | 10 | | [44] |
| Background infection rate in community outside shelter | $i_a$ | | | |
| Seattle A | | 561 infections/1,000,000 person-days | | [38] |
| Seattle B | | 543 infections/1,000,000 person-days | | [38] |
| Seattle C | | 543 infections/1,000,000 person-days | | [38] |
| Boston | | 2018 infections/1,000,000 person-days | | [39] |
| San Francisco | | 445 infections/1,000,000 person-days | | [40] |
| Relative risk of infection for homeless individuals | $r$ | 2 | | |
| Background infection rate in homeless community outside shelter | $\epsilon = ri_a$ | | | |
| *Intervention scenarios* | | | | |
| Simulation duration | | 30 days | | Assumed |
| Number of residents | | 250 | | Assumed |
| Number of staff | | 50 | | Assumed |
| Initial number of infected individuals | | 1 (assumed latent) | | Assumed |
| Age- and co-morbidity stratification | | Same as for San Francisco shelter | | Assumed |
| Basic reproduction number | $R_0$ | | | |
| “low-risk” | | 1.5 | | Assumed |

| | | | | |
|---|---|---|---|---|
| “Seattle” | | 2.9 | | Calibrated |
| “Boston” | | 3.9 | | Calibrated |
| “San Francisco” | | 6.2 | | Calibrated |
| Infection-to-reported-case ratio | $\gamma$ | 4 | | [45] |
| Background infection rate in community outside shelter | $i_a$ | | 0–439 | |
| No background infection | | 0 infections/1,000,000 person-days | | |
| Low | | 122 infections/1,000,000 person-days | | [39] |
| Moderate | | 260 infections/1,000,000 person-days | | [38–40] |
| High | | 439 infections/1,000,000 person-days | | [40] |
| Symptom screening | | | | |
| Sensitivity | | 0.4 | 0.3–0.5 | Assumed based on [65] |
| Specificity | | 0.9 | 0.8–0.9 | Assumed |
| Compliance of symptomatic individuals with PCR testing | | 80% | 50–100% | Assumed |
| PCR testing | | | | |
| Sensitivity | | 0.75 | 0.6–0.9 | [17–20] |
| Specificity | | 1 | 0.95–1 | [17,20] |
| Frequency | | Twice weekly | Daily–Monthly | [66–68] |
| Compliance | | 80% | 50–100% | Assumed |
| Masks | | | | |
| Effectiveness at reducing infectious material exhaled | $e_{ex}$ | 30% | 10-50% | [25,26,31] |
| Effectiveness at reducing infectious material inhaled | $e_{in}$ | 40% | 20-60% | [25,26,31] |
| Compliance | $c$ | 60% | 30–100% | [26,37] |

**Table S6. Different intervention strategies tested**

| Strategy | Interventions | | | | |
|---|---|---|---|---|---|
| | **Daily symptom screening** | **Twice-weekly PCR testing of residents** | **Twice-weekly PCR testing of staff** | **Universal masking** | **Relocation of high-risk individuals** |
| (1) Symptom screening | ✓ | ✗ | ✗ | ✗ | ✗ |
| (2) Routine PCR testing | ✓ | ✓ | ✓ | ✗ | ✗ |
| (3) Universal mask wearing | ✓ | ✗ | ✗ | ✓ | ✗ |
| (4) Relocation of high-risk individuals | ✓ | ✗ | ✗ | ✗ | ✓ |
| (5) Routine PCR testing of staff only | ✓ | ✗ | ✓ | ✗ | ✗ |
| (6) Combination strategy | ✓ | ✓ | ✓ | ✓ | ✓ |

### Table S7. Estimated epidemiologic parameters based on observed outbreak data from homeless shelters in Seattle, Boston and San Francisco

| Shelter | Basic reproduction number $R_0$, median (95% CI)* | Number of latently infected individuals who initially entered shelter $E_0$, median (95% CI)* | Number of days prior to first reported infection that infected individuals entered shelter† $D$, median (95% CI)* |
|---|---|---|---|
| Seattle A | 2.9 (1.1–7.3) | 3 (1–5) | 16 (9–25) |
| Seattle B | 2.9 (1.1–6.7) | 3 (1–5) | 10 (7–14) |
| Seattle C | 3.0 (1.2–7.2) | 3 (1–5) | 15 (8–24) |
| Boston | 3.9 (2.2–7.6) | 3 (1–5) | 15 (9–23) |
| San Francisco | 6.2 (4.0–7.9) | 3 (1–5) | 21 (17–26) |

CI = credible interval.

Data was available for three shelters in Seattle (labeled A-C).

* 95% CIs calculated as 2.5%–97.5% quantile interval of posterior distribution.

† $D$ is calculated from the estimated time since introduction of infection into the shelter at the end of data collection, $T$, as:

$D = \text{date first case identified} - (\text{end date of data collection} - T) + 1$

### Table S8. Estimated cumulative infection incidence at the end of the PCR testing period in homeless shelters in Seattle, Boston and San Francisco

| Shelter | Cumulative infection incidence, median (95% CI)*, % |
|---|---|
| Seattle A | 40 (10–81) |
| Seattle B | 14 (1–41) |
| Seattle C | 37 (12–71) |
| Boston | 64 (52–78) |
| San Francisco | 83 (72–92) |

CI = credible interval.

* 95% CIs calculated as 2.5%–97.5% quantile interval of posterior distribution.

**Table S9. Probability of averting an outbreak over a 30-day period in a generalized homeless shelter* with simulated infection control strategies for different background infection rates in the community outside the shelter**

| Infection control strategy | Probability of averting an outbreak $R_0 = 1.5$ (low-risk) | $R_0 = 2.9$ (Seattle) | $R_0 = 3.9$ (Boston) | $R_0 = 6.2$ (San Francisco) |
|---|---|---|---|---|
| Background infection rate = 0 | | | | |
| (1) Symptom screening | 0.63 | 0.31 | 0.24 | 0.13 |
| (2) Routine twice-weekly PCR testing | 0.78 | 0.42 | 0.33 | 0.13 |
| (3) Universal mask wearing | 0.81 | 0.54 | 0.43 | 0.25 |
| (4) Relocation of high-risk individuals | 0.63 | 0.31 | 0.23 | 0.11 |
| (5) Routine twice-weekly PCR testing of staff only | 0.66 | 0.35 | 0.25 | 0.14 |
| (6) Combination strategy | 0.91 | 0.68 | 0.55 | 0.27 |
| Background infection rate = 260/1,000,000/day | | | | |
| (1) Symptom screening | 0.17 | 0.04 | 0.02 | 0.01 |
| (2) Routine twice-weekly PCR testing | 0.35 | 0.07 | 0.06 | 0.01 |
| (3) Universal mask wearing | 0.41 | 0.13 | 0.07 | 0.02 |
| (4) Relocation of high-risk individuals | 0.16 | 0.04 | 0.02 | 0.01 |
| (5) Routine twice-weekly PCR testing of staff only | 0.21 | 0.04 | 0.02 | 0.00 |
| (6) Combination strategy | 0.60 | 0.25 | 0.16 | 0.03 |
| Background infection rate = 439/1,000,000/day | | | | |
| (1) Symptom screening | 0.08 | 0.01 | 0.01 | 0.00 |
| (2) Routine twice-weekly PCR testing | 0.19 | 0.03 | 0.01 | 0.00 |
| (3) Universal mask wearing | 0.23 | 0.04 | 0.02 | 0.01 |
| (4) Relocation of high-risk individuals | 0.06 | 0.01 | 0.01 | 0.00 |
| (5) Routine twice-weekly PCR testing of staff only | 0.09 | 0.01 | 0.00 | 0.00 |
| (6) Combination strategy | 0.41 | 0.12 | 0.05 | 0.02 |

$R_0$ = basic reproduction number.
* Generalized homeless shelter defined as 250 residents and 50 staff.

**Table S10. Reductions in the total number of infections and symptomatic cases over a 30-day period in a generalized homeless shelter* with simulated infection control strategies for different background infection rates in the community outside the shelter**

| Infection control strategy | Median reduction in total infections (%) | | | | Median reduction in total symptomatic cases (%) | | | |
|---|---|---|---|---|---|---|---|---|
| | $R_0 = 1.5$ (low-risk) | $R_0 = 2.9$ (Seattle) | $R_0 = 3.9$ (Boston) | $R_0 = 6.2$ (San Francisco) | $R_0 = 1.5$ (low-risk) | $R_0 = 2.9$ (Seattle) | $R_0 = 3.9$ (Boston) | $R_0 = 6.2$ (San Francisco) |
| Background infection rate = 0 | | | | | | | | |
| (1) Symptom screening | 55 | 60 | 52 | 13 | 33 | 56 | 49 | 28 |
| (2) Routine PCR testing | 88 | 85 | 82 | 40 | 60 | 75 | 75 | 54 |
| (3) Universal mask wearing | 86 | 94 | 94 | 80 | 67 | 86 | 87 | 84 |
| (4) Relocation of high-risk individuals | 53 | 60 | 48 | 10 | 50 | 60 | 51 | 32 |
| (5) Routine PCR testing of staff only | 72 | 65 | 56 | 18 | 50 | 59 | 53 | 36 |
| (6) Combination strategy | 96 | 97 | 98 | 92 | 75 | 92 | 94 | 93 |
| Background infection rate = 122/1,000,000/day | | | | | | | | |
| (1) Symptom screening | 40 | 45 | 38 | 7 | 33 | 39 | 41 | 24 |
| (2) Routine PCR testing | 61 | 75 | 69 | 31 | 50 | 63 | 65 | 54 |
| (3) Universal mask wearing | 67 | 83 | 86 | 69 | 50 | 76 | 81 | 79 |
| (4) Relocation of high-risk individuals | 36 | 44 | 34 | 6 | 42 | 44 | 45 | 32 |
| (5) Routine PCR testing of staff only | 50 | 51 | 45 | 8 | 33 | 48 | 46 | 28 |
| (6) Combination strategy | 72 | 90 | 92 | 88 | 67 | 85 | 89 | 92 |
| Background infection rate = 260/1,000,000/day | | | | | | | | |
| (1) Symptom screening | 33 | 37 | 32 | 5 | 25 | 32 | 34 | 17 |
| (2) Routine PCR testing | 53 | 70 | 69 | 22 | 43 | 60 | 67 | 40 |
| (3) Universal mask wearing | 59 | 76 | 79 | 63 | 50 | 69 | 77 | 73 |
| (4) Relocation of high-risk individuals | 30 | 36 | 31 | 5 | 35 | 40 | 41 | 27 |
| (5) Routine PCR testing of staff only | 40 | 42 | 38 | 7 | 29 | 38 | 41 | 21 |
| (6) Combination strategy | 66 | 86 | 90 | 79 | 63 | 81 | 88 | 85 |
| Background infection rate = 439/1,000,000/day | | | | | | | | |
| (1) Symptom screening | 31 | 36 | 25 | 3 | 23 | 34 | 31 | 16 |
| (2) Routine PCR testing | 50 | 67 | 60 | 20 | 42 | 61 | 61 | 43 |
| (3) Universal mask wearing | 56 | 74 | 75 | 53 | 48 | 68 | 73 | 67 |

| | | | | | | | | |
|---|---|---|---|---|---|---|---|---|
| (4) Relocation of high-risk individuals | 28 | 35 | 24 | 3 | 33 | 42 | 37 | 25 |
| (5) Routine PCR testing of staff only | 36 | 41 | 33 | 4 | 27 | 38 | 37 | 18 |
| (6) Combination strategy | 63 | 84 | 86 | 75 | 62 | 80 | 85 | 83 |

$R_0$ = basic reproduction number.
* Generalized homeless shelter defined as 250 residents and 50 staff.

### Table S11. Numbers of PCR tests used under each infection control strategy

| Infection control strategy | Mean number of tests used per person per month* |
|---|---|
| (1) Symptom screening | 2.0 |
| (2) Routine PCR testing | 6.6 |
| (3) Universal mask wearing | 2.0 |
| (4) Relocation of high-risk individuals | 2.0 |
| (5) Routine PCR testing of staff only | 2.8 |
| (6) Combination strategy | 6.6 |

* All strategies use tests as they all include daily symptom screening.

Numbers only shown for $R_0 = 2.9$ (Seattle), chosen as representative example, and a background infection rate of 122/1,000,000/day as numbers vary little with $R_0$ and background rate.

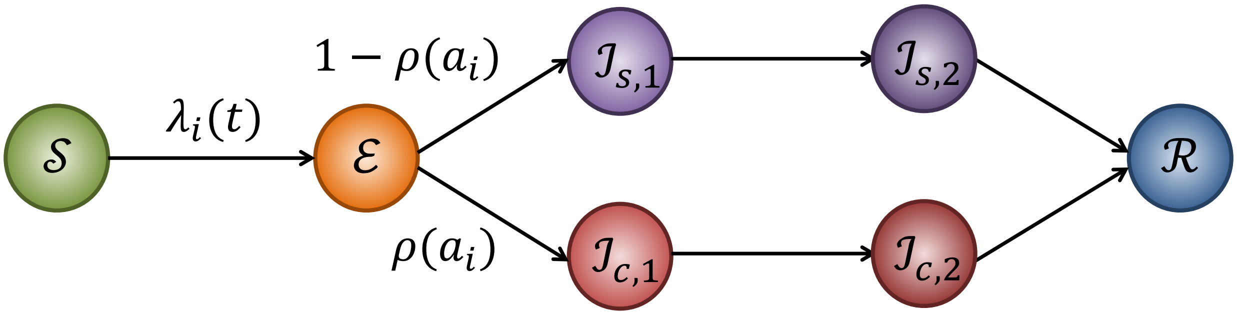


**Figure S1. Structure of stochastic individual-level susceptible-exposed-infectious-recovered ($\mathcal{S}$-$\mathcal{E}$-$\mathcal{I}$-$\mathcal{R}$) model of COVID-19 transmission in homeless shelter.** Notation as defined in Tables S1 and S5 and Equation (1).

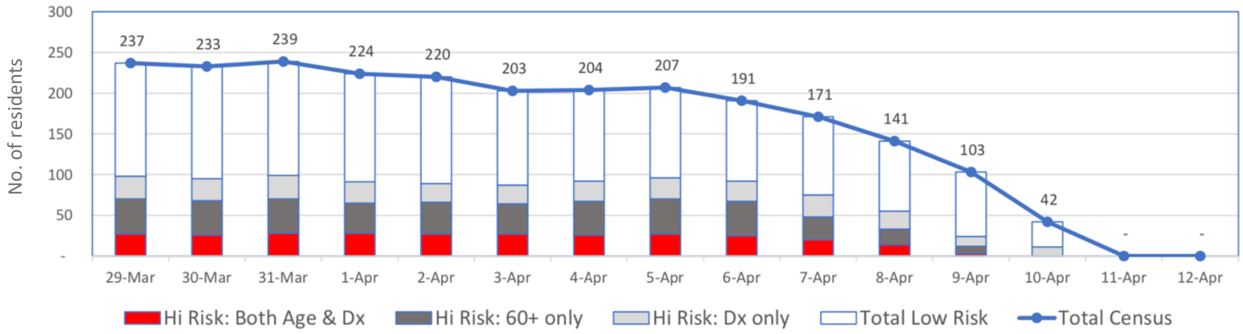


**Figure S2. Daily numbers of residents by risk group present in the San Francisco shelter March 29–April 10, 2020.** Shelter was disbanded April 11. Dx = co-morbidity.

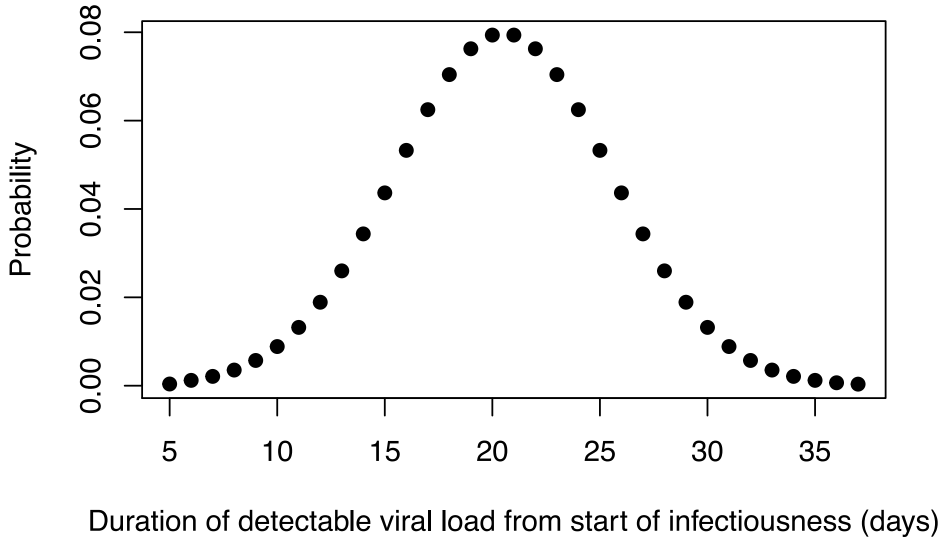


**Figure S3. Distribution of duration of detectable viral load from start of late infectious stage**

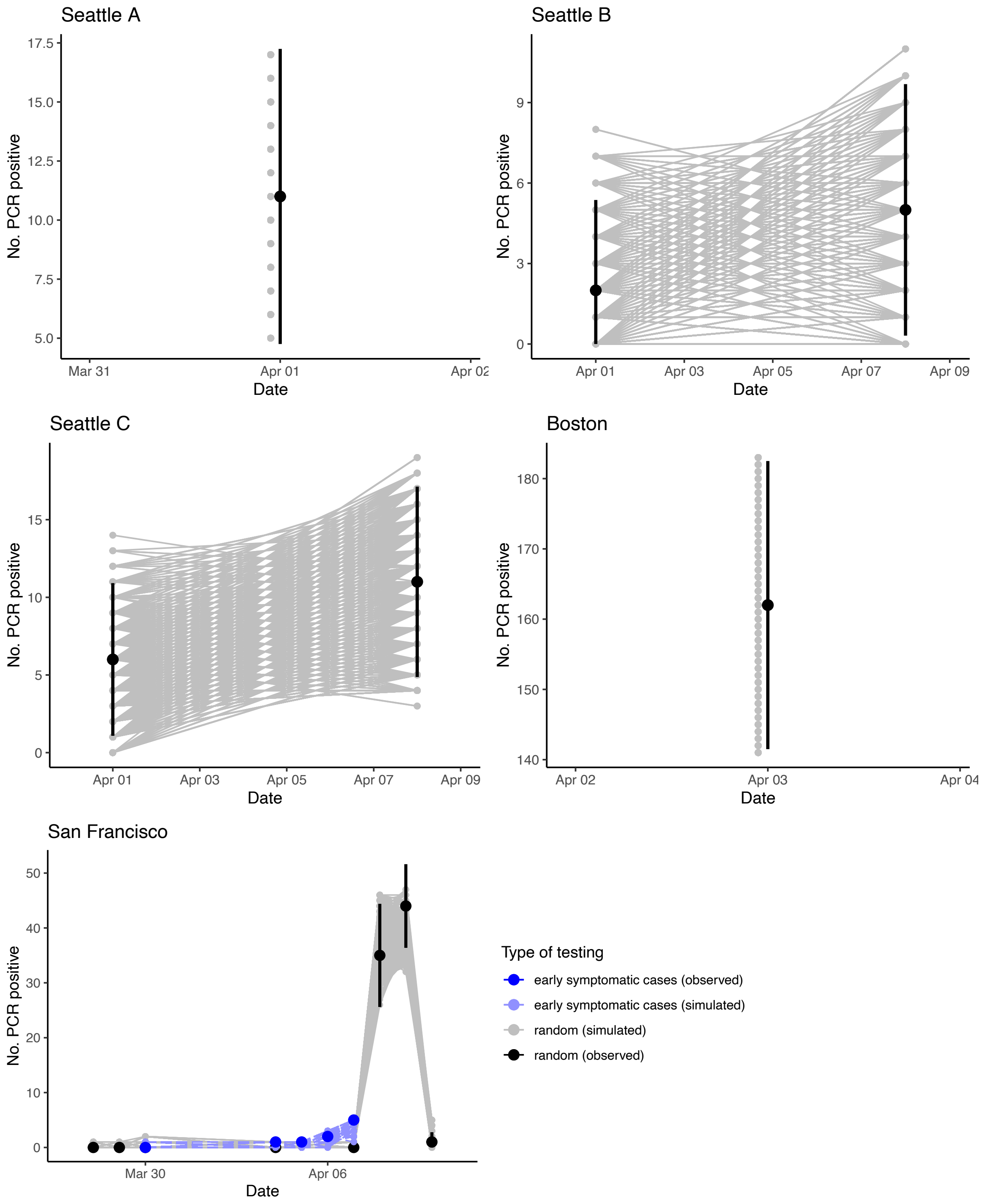


**Figure S4. Calibration of microsimulation to observed PCR testing data from outbreaks in homeless shelters in Seattle, Boston and San Francisco.** Data was available for three shelters in Seattle (labeled A-C). Vertical black lines show exact binomial 95% confidence intervals for observed numbers of PCR-positive individuals in random testing.

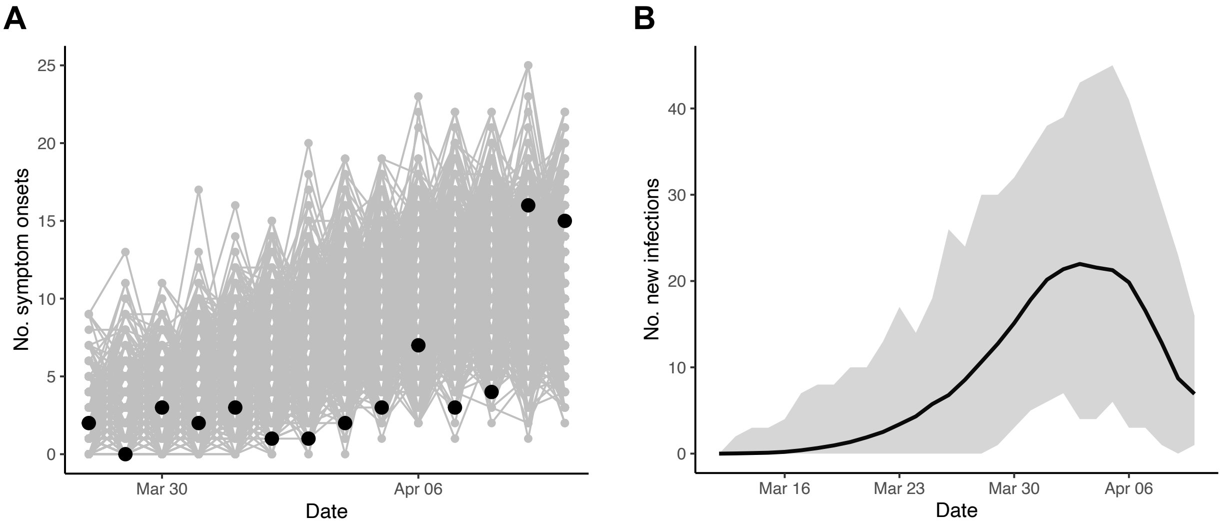


**Figure S5. Calibration of microsimulation to additional data from San Francisco shelter outbreak.** (A) Calibration to daily numbers of symptom onsets. Black dots show reported number of symptom onsets, grey lines show simulation output. (B) Estimated daily numbers of new infections over time in 1000 calibrated simulations. Black line and grey shaded region show mean and range respectively across all simulations.

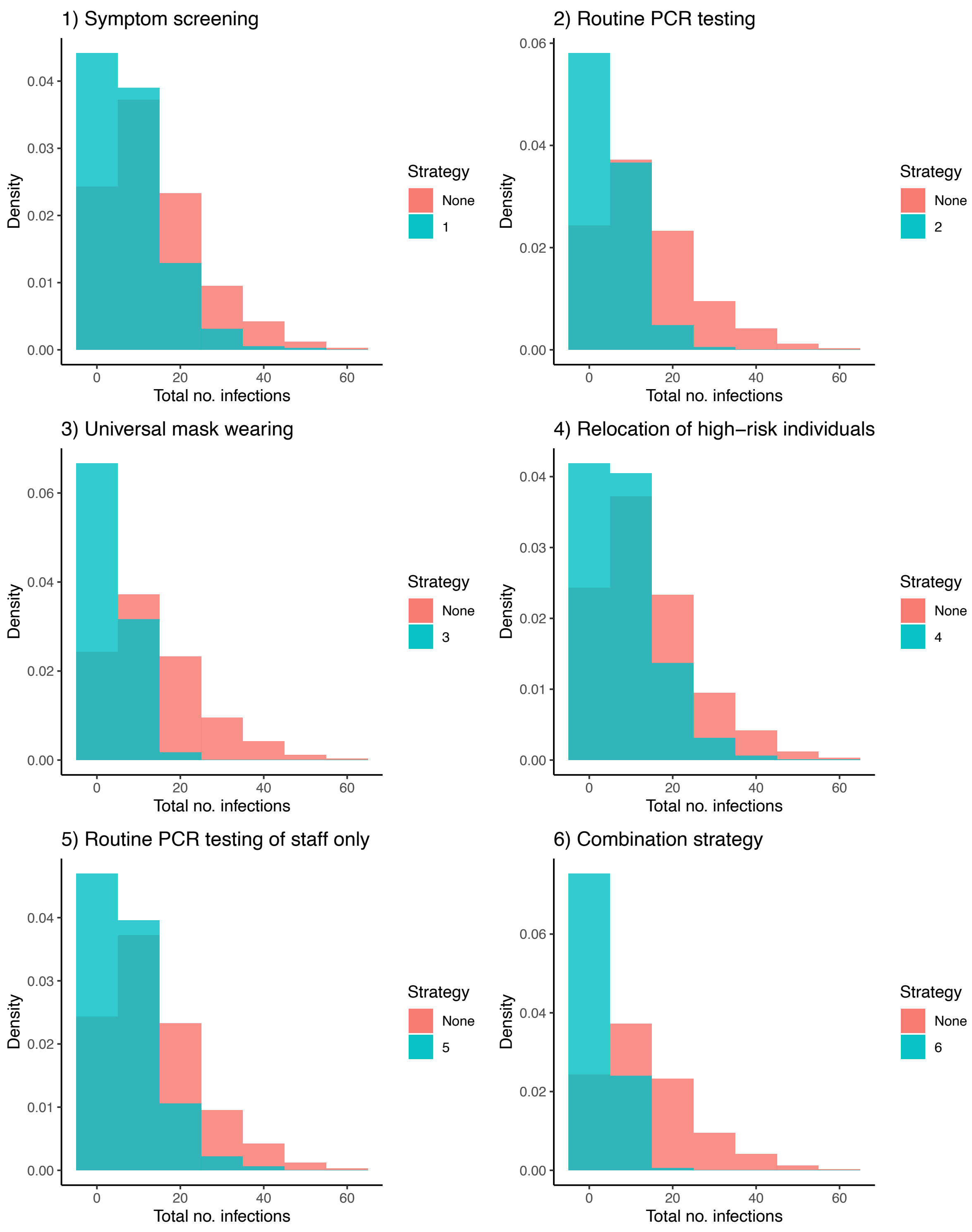


**Figure S6. Outbreak size distributions 30 days after introduction of infection in a generalized homeless shelter under different infection control strategies for $R_0 = 1.5$ (low-risk setting).** Red and green histograms show outbreak size distributions with no interventions and under the intervention strategy respectively. Background infection rate of 122/1,000,000 person-days.

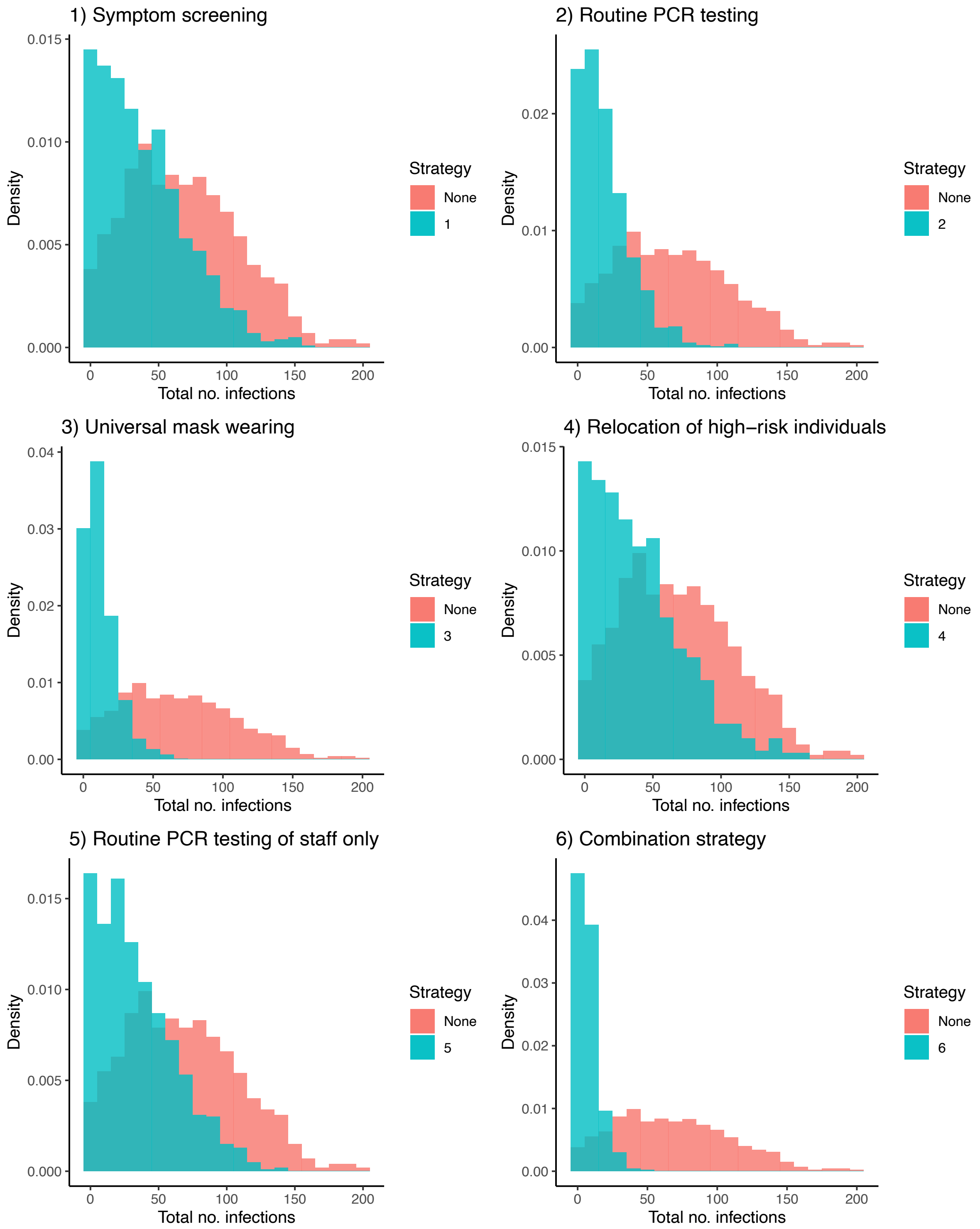


**Figure S7. Outbreak size distributions 30 days after introduction of infection in a generalized homeless shelter under different infection control strategies for $R_0 = 2.9$ (Seattle).** Red and green histograms show outbreak size distributions with no interventions and under the intervention strategy respectively. Background infection rate of 122/1,000,000 person-days.

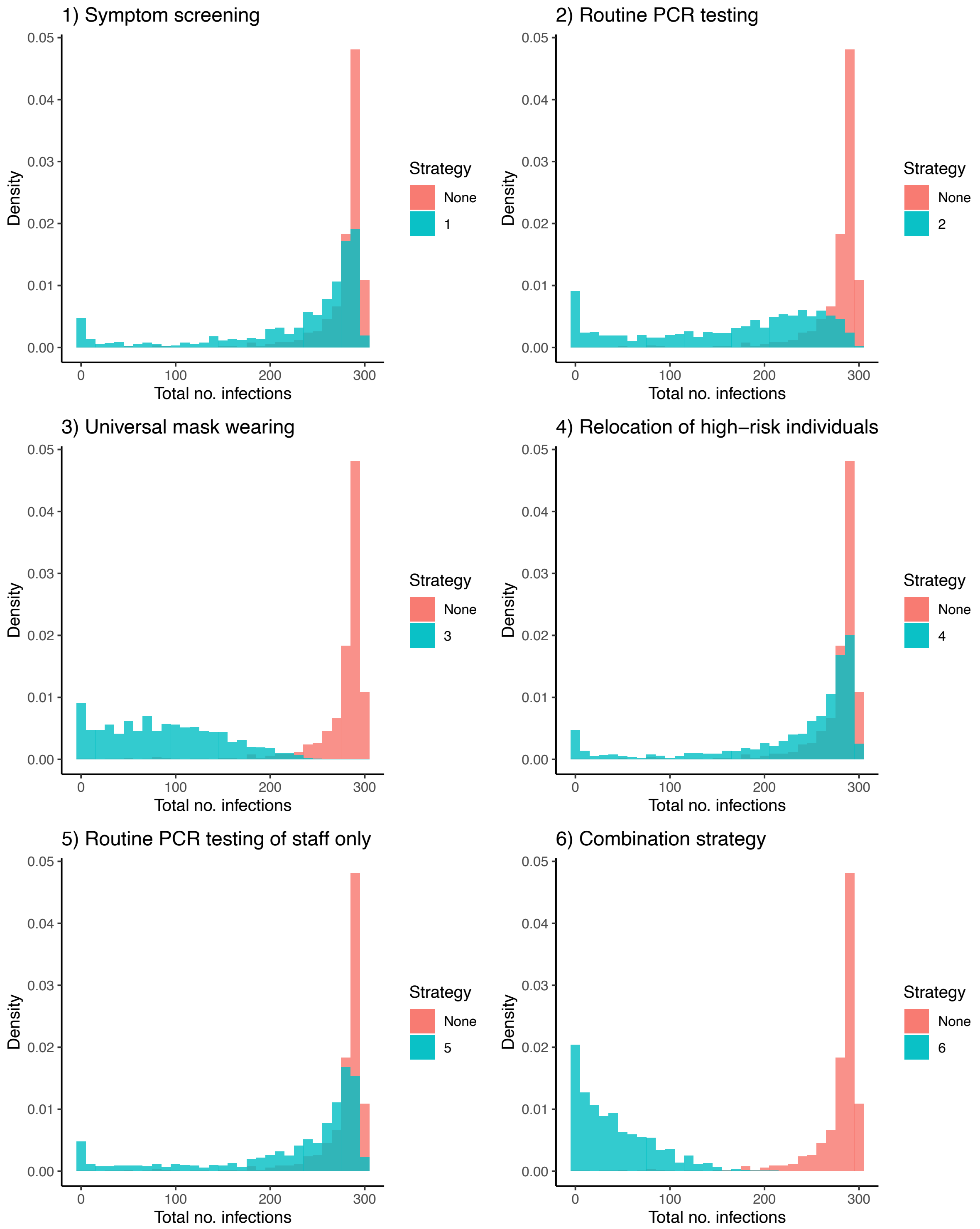


**Figure S8. Outbreak size distributions 30 days after introduction of infection in a generalized homeless shelter under different infection control strategies for $R_0 = 6.2$ (San Francisco).** Red and green histograms show outbreak size distributions with no interventions and under the intervention strategy respectively. Background infection rate of 122/1,000,000 person-days.

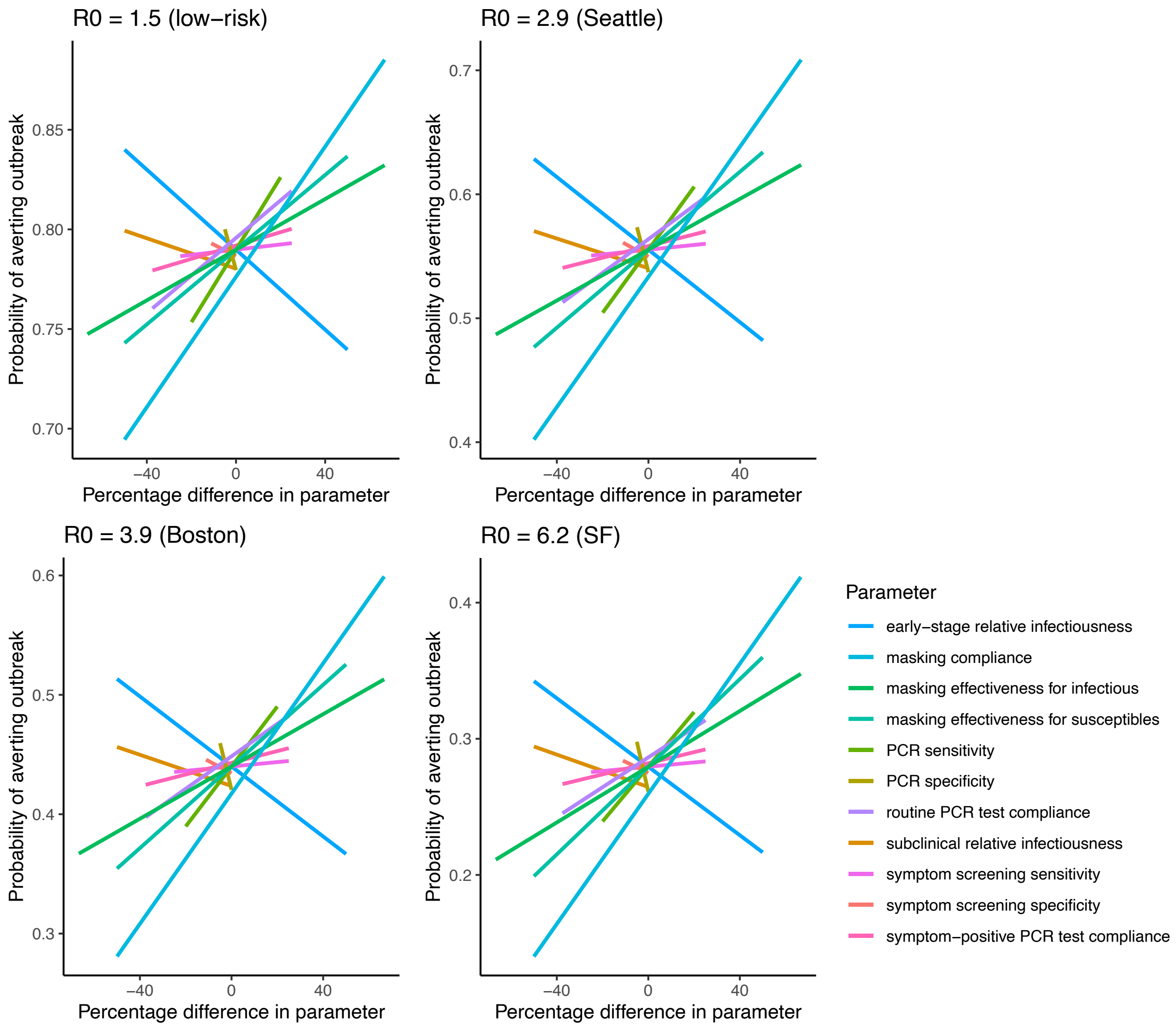


**Figure S9. Spider diagrams showing the sensitivity of the estimated probability of averting an outbreak to variation in key natural history and intervention parameters for different $R_0$ values**. The horizontal axis shows the percentage variation in each parameter relative to its base case value (see Table S5). The vertical axis shows the mean probability of averting an outbreak for each parameter under the combination strategy across all combinations of the minimums and maximums of the other parameter values.

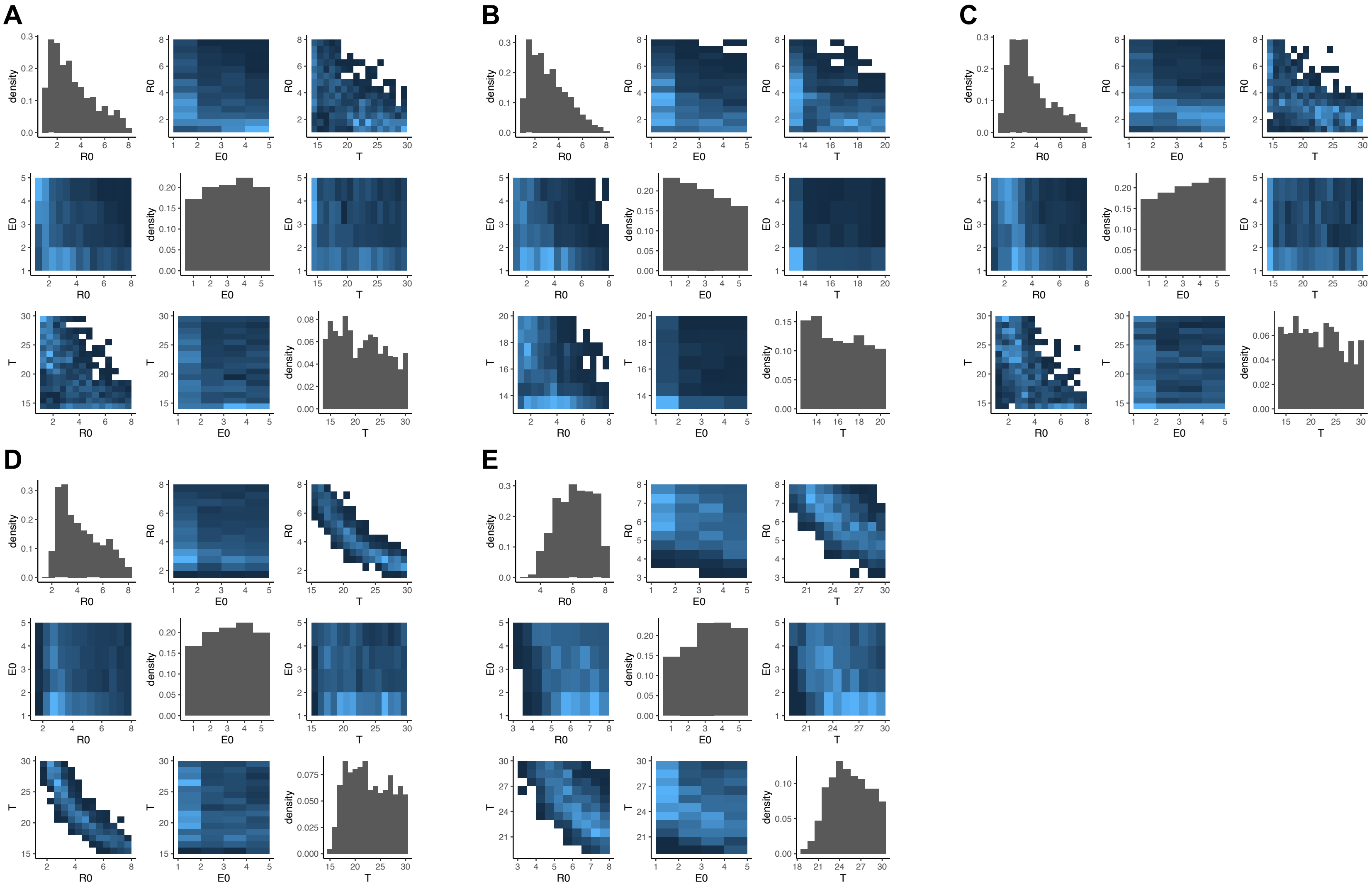


**Figure S10. Posterior distributions and pairwise correlation plots for calibrated model parameters – $R_0$, $E_0$ and $T$ – for (A)-(C) Seattle shelters A–C, (D) Boston shelter and (E) San Francisco shelter.** $R_0$ = basic reproduction number; $E_0$ = number of latently infected individuals who initially entered the shelter; $T$ = number of days before the end of data collection that these individuals entered the shelter.
